# Estimating the Case Fatality Ratio for COVID-19 using a Time-Shifted Distribution Analysis

**DOI:** 10.1101/2020.10.25.20216671

**Authors:** B. S. Thomas, N. A. Marks

## Abstract

Estimating the case fatality ratio (CFR) for COVID-19 is an important aspect of public health. However, calculating CFR accurately is problematic early in a novel disease outbreak, due to uncertainties regarding the time course of disease and difficulties in diagnosis and reporting of cases. In this work, we present a simple method for calculating the case fatality ratio using only public case and death data over time by exploiting the correspondence between the time distributions of cases and deaths. The time-shifted distribution (TSD) analysis generates two parameters of interest: the delay time between reporting of cases and deaths and the case fatality ratio. These parameters converge reliably over time once the exponential growth phase has finished. Analysis is performed for early COVID-19 outbreaks in many countries, and we discuss corrections to CFR values using excess-death and seroprevalence data to estimate the infection fatality ratio (IFR). While CFR values range from 0.2-20% in different countries, estimates for IFR are mostly around 0.5-0.8% for countries that experienced moderate outbreaks and 1-3% for severe outbreaks. The simplicity and transparency of TSD analysis enhance its usefulness in characterizing a new disease as well as the state of the health and reporting systems.

## Introduction

The novel coronavirus SARS-CoV-2, and its attendant disease, COVID-19, first appeared in late 2019 in Wuhan, China. Since then, studies and estimates of the transmissibility and virulence of COVID-19 have abounded, with widely varying results [1-6]. Virulence is often measured using the case fatality ratio (also called case fatality rate or case fatality risk, CFR), which is the number of deaths due to a disease as a proportion of the number of people diagnosed with the disease. The CFR is dependent on the particular pathogen (and its mechanism of action) and the immune response of the host, which can depend on age, sex, genetic factors and pre-existing medical conditions. Environmental factors such as climate and health system may also affect CFR. It is important to accurately quantify the CFR of a new disease to inform policy, communication and public health measures.

Calculating the case fatality ratio requires data on cases and deaths over time, either for individuals or populations. In general, the CFR is based on diagnosed cases of disease rather than the number of actual infections (which is difficult to measure); there may be many more infections than reported cases, depending on the expression of symptoms and the degree of testing. The simplest estimate of CFR is to divide the cumulative number of deaths by the cumulative number of cases at a given time, known as the crude (or naïve) CFR. However, the crude CFR tends to underestimate the CFR during an outbreak because at any given time, some of the existing known cases will prove fatal and need to be included in the death count. This bias is known as right-censoring and obscures the CFR of a new disease early in the course of the outbreak, particularly before the time course of the disease is characterised. Further, even once the distribution of times from onset of disease to death is known, it can be difficult to use this information to accurately correct the crude CFR. An alternative method is to use data for closed cases only, once patients have recovered or died (eg [7, 8]), yet this information is also difficult to obtain during an outbreak and may be biased towards a particular demographic or skewed by delays in reporting of recoveries. Other biases in calculating CFR include under-ascertainment of mild or asymptomatic cases, time lags in testing and reporting, and the effects of intervention approach, demographics and reporting schemes [9].

There are many published calculations of case fatality ratios for COVID-19 using various datasets from different countries and using a range of methods. In some places, initial outbreaks have now concluded and the final crude CFR accurately reflects the overall ratio of reported deaths to cases. In many other places, outbreaks are continuing. Questions remain regarding quality of data, methods of calculation and even the possibility of changes in the CFR over time. These continuing uncertainties make it necessary to improve estimates of the CFR by refining the methods used to calculate it. In essence, this means finding the best way to correct the crude CFR for biases due to time lags and other factors. Most previously published studies make use of a parametrised distribution of times from onset (or hospitalisation) to death, determined from individual case data from early in the outbreak (largely from China) [5, 10-12], which is then used in combination with statistical methods to estimate the CFR using population-level data on cases and deaths [5, 10, 11, 13]. Various assumptions are made in these analyses, including the form (and transferability) of the time course of cases, time lags in reporting or testing or hospitalisation, and estimates of the proportion of cases being detected. Early values of CFR obtained using these methods range from 1-18%, with the highest values obtained for China: 4-18% early in the outbreak [5, 10], 12% in Wuhan and as low as 1% outside Hubei province [11]. Values reported outside China include 1-5% for early cases in travellers [5, 10], and 1-4% in Korea [13]. Case fatality ratios have also been shown to vary greatly with the age of the patient [5], which limits the transferability of parameters based on case studies. The specific data requirements and the range of approximations and assumptions required by statistical methods can make it difficult to interpret or rely on the results of such analyses, since biases can be obscured.

### Time-shifted distribution analysis for COVID-19 data

The time-shifted distribution (TSD) analysis method began with an observation that the shape of the evolving time distribution of COVID-19 cases in a given country often closely matches the shape of the corresponding distribution of COVID-19 deaths — simply shifted by a number of days and linearly scaled in magnitude. This is illustrated in Figure 1 for COVID-19 cases and deaths in Italy (data from [14], 3-day averaged data shown); the time-shifted relationship between case and death distributions can be seen in both cumulative and daily tallies. We can understand this shift from the perspective of the time delay between diagnosis and death or recovery. However, the closeness of the match reflects a much simpler apparent relationship than that suggested or assumed by conventional analyses, which relate deaths and cases using statistical parametric models that incorporate a broad distribution of expected times between diagnosis (or onset) and death, usually generated from case study data (e.g. [15]).

**Figure 1:**
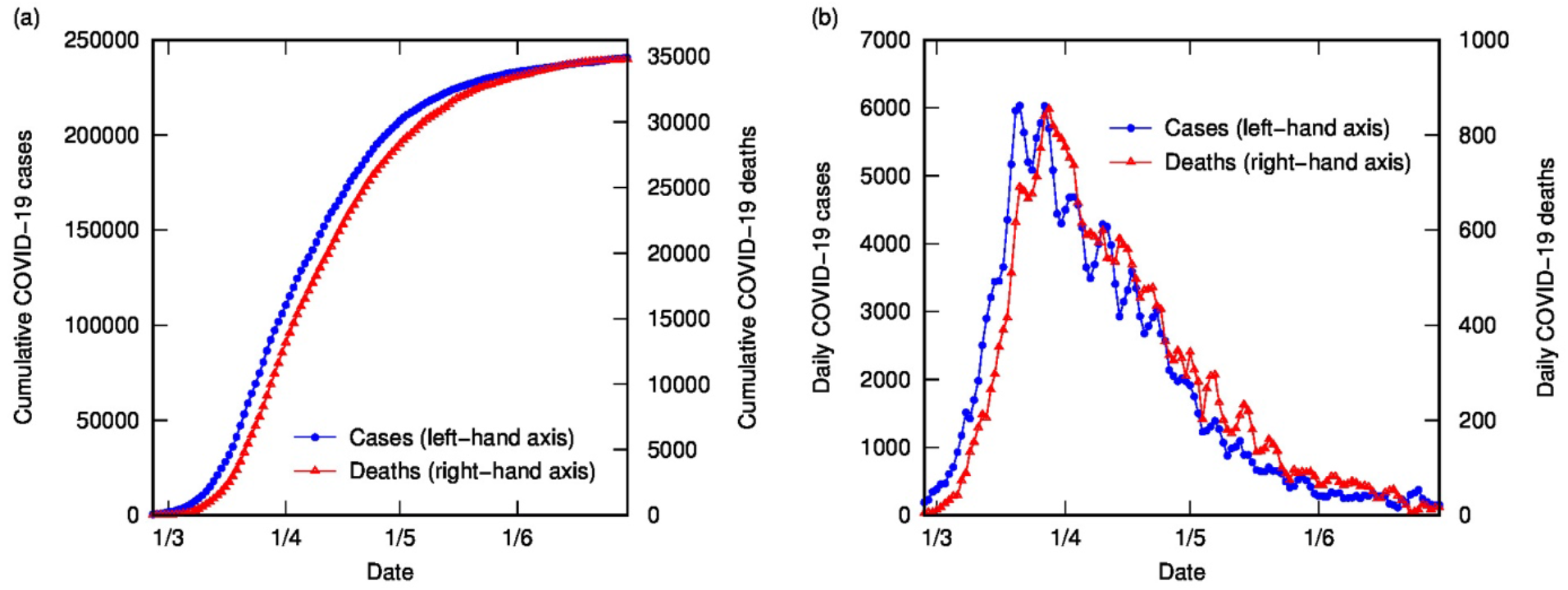
COVID-19 cases and deaths in Italy to end of June (2020), using 3-day-averaged data: (a) Cumulative cases (left-hand axis) and deaths (right-hand axis); (b) Daily cases (left-hand axis) and deaths (right-hand axis).

This observation suggests that there are two parameters of interest: the number of days separating the case and death distributions (called the delay time or *t*_*d*_), and the scaling factor between the time-shifted case data and the death data, *λ*. For the optimal value of *t*_*d*_, there is a simple linear relationship between cumulative number of deaths at time *t, D(t)*, and cumulative number of cases at time *t − t*_*d*_, *C(t − t*_*d*_), with gradient *λ*:

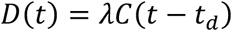

To find the optimal value for *t*_*d*_, we test integer values from zero to 25 days. For each value of *t*_*d*_, we plot *D(t)* as a function of *C(t − t*_*d*_) (for all *t*) and perform a linear regression using Matlab. The value of *t*_*d*_ is chosen on the basis of the lowest root-mean-squared error in the linear regression analysis and the value of *λ* is the gradient of the corresponding line.

Figure 2 contains the results of this analysis for Italy. Figure 2 (a) shows the error from linear regression of *D*(*t*) versus *C(t − t*_*d*_) as a function of delay time, with a clear minimum at four days. Figure 2 (b) shows *D*(*t*) versus *C(t − t*_*d*_) for different delay times: the optimal value of four days (with linear fit shown) as well as some other representative values, displaying the convergence of nonlinear to linear relationship with optimized *t*_*d*_. Figure 2 (c) and (d) show the excellent correlation of time-shifted and scaled case data and death data (cumulative and daily, respectively), using a delay time of four days and a linear scaling factor of 0.144. What do these parameters represent? The delay time is presumably a measure of the delay between reporting of confirmed cases and reporting of COVID-19-related deaths. While four days seems very short compared to current estimates of the mean delay between onset of COVID-19 symptoms and death (or even between hospitalisation and death), which is around 12-22 days with a large variance [2, 5, 7, 10, 16], it is possible to rationalise the shorter apparent delay on the basis of delays in testing, diagnosis and reporting of the disease, particularly in countries where the outbreak is severe. For example, in Italy from late February, testing was prioritized for “patients with more severe clinical symptoms who were suspected of having COVID-19 and required hospitalization” [17]; a subsequent delay in test results could account for the rather short delay between reported diagnosis and death. This shows the inherent danger in analysing such datasets using time-delay distributions from specific case data (which presumes a much longer delay time). Moreover, the delay time may provide some useful information about relative conditions in various countries.

**Figure 2:**
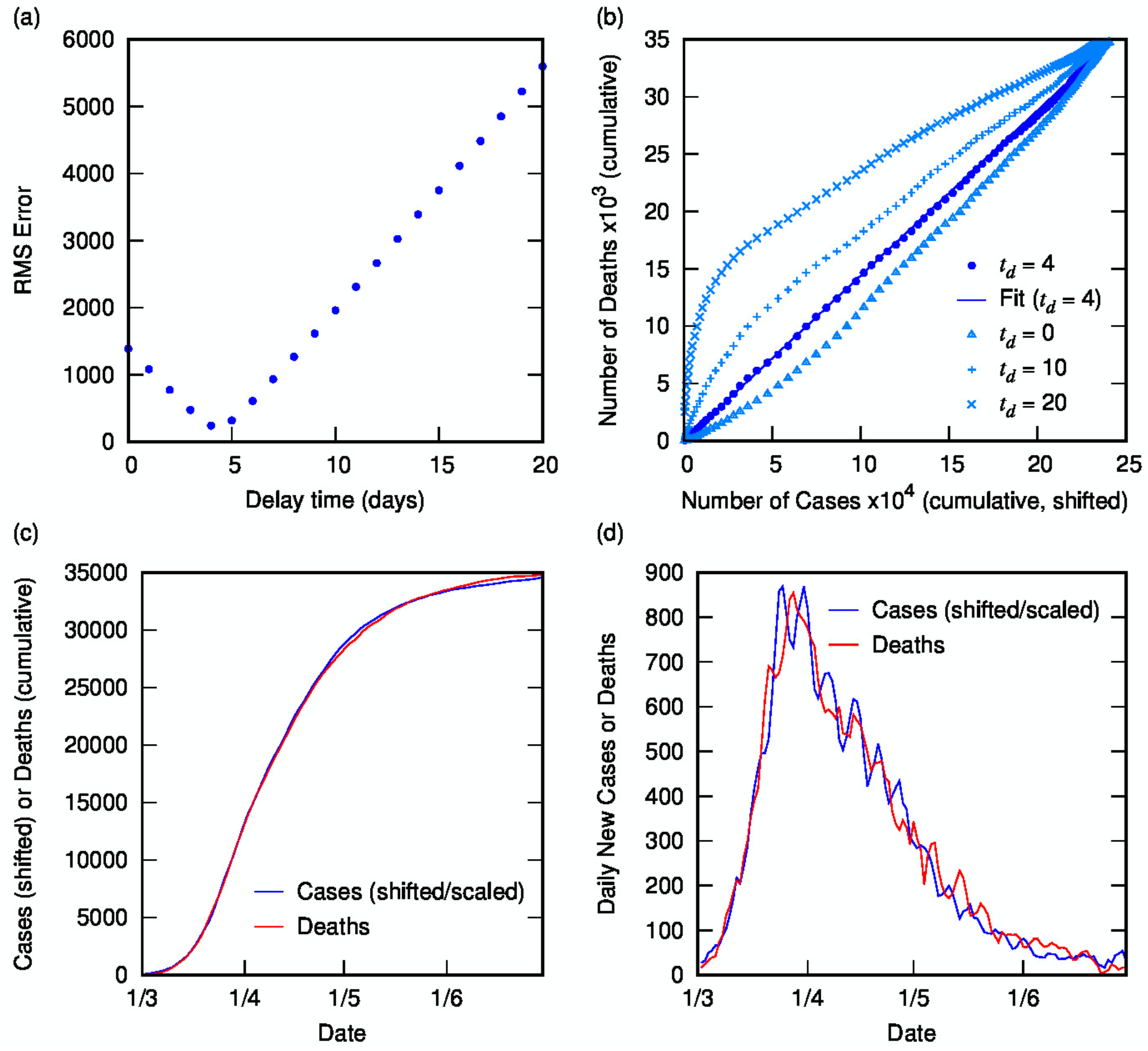
Time-shifted distribution analysis for Italy: (a) Root-mean-squared error in linear regression as a function of delay time, *t*_*d*_; (b) Cumulative deaths as a function of cumulative cases, time-shifted by various *t*_*d*_ values, including the optimal value of 4 days with linear regression shown; (c) Overlay of cumulative deaths and time-shifted (and scaled) cases as a function of time, using optimal *t*_*d*_; (d) Overlay of daily deaths and time-shifted (and scaled) cases as a function of time using optimal *t*_*d*_.

Using a time delay of four days in Italy, the scaling factor of 0.144 represents the ratio of deaths to cases, or in other words, an estimate for the case fatality ratio, converging towards the crude CFR with time. The calculated CFR of 14.4% is almost identical to the crude CFR of 14.5% at the end of June, which is a good estimate for the “true” CFR at the end of the outbreak. An interesting question is, at what point in the outbreak does the CFR calculated using the TSD analysis give a good approximation to the final value? This is important because early estimates of CFR are vital for informing public health decisions. Figure 3 shows the CFR calculated at various stages of the outbreak using data available to that point. Errors represent uncertainty in the linear regression as well as in *t*_*d*_. The earliest data predict a longer delay time, which results in a higher predicted CFR; once the value of *t*_*d*_ has stabilised (from 26 March), the predicted value of CFR is also very stable, and also remarkably accurate (14.4%), compared to the crude value of 10.3% at that time. Even a week earlier, the calculated CFR of 16.6% is a better estimate than the crude estimate of 8.4%.

**Figure 3:**
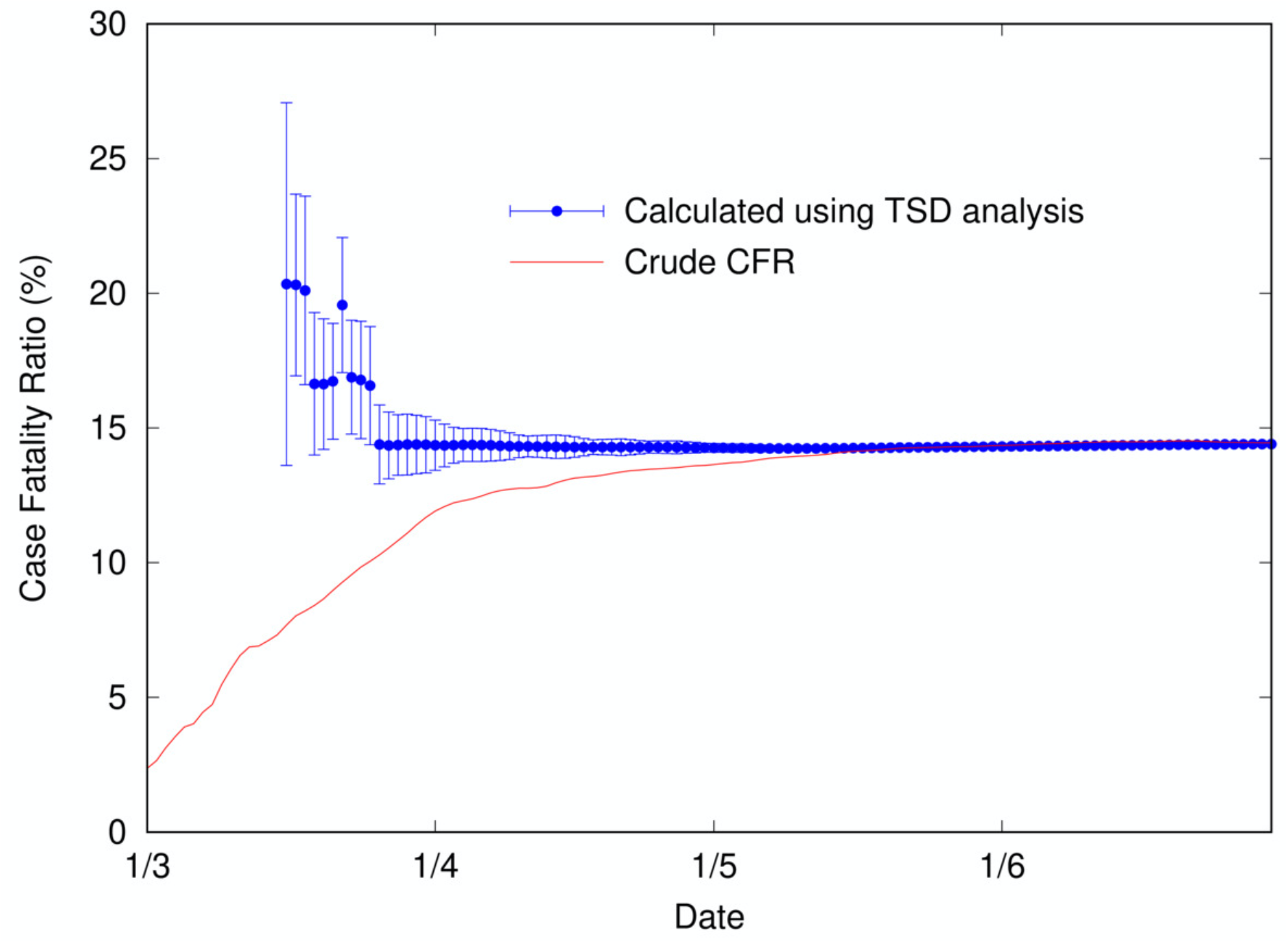
Calculated case fatality ratio (using TSD analysis) for COVID-19 in Italy (2020) as a function of time during an outbreak, alongside the crude CFR.

It appears that this simple analysis generates two parameters of significant interest: the apparent delay between reporting of related cases and deaths, and the CFR. The estimates of these parameters (which can be determined unequivocally once an outbreak is concluded) can be calculated during the course of an outbreak and give a better approximation than the crude CFR. It should be noted that such an analysis cannot be applied during purely exponential growth, because time-shifting (horizontally) and scaling (vertically) an exponential function are equivalent operations, as: 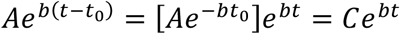, which means that any value of *t*_*d*_ will give an equivalent relationship between *C(t − t*_*d*_) and *D*(*t*) with gradient depending on *t*_*d*_. Therefore, the TSD analysis is only valid once exponential growth ends and the daily case rate is approaching (or past) its peak. Alternatively, an estimate for *t*_*d*_ could be used, but this reduces the simplicity and transparency of the model. We note that others have calculated an “adjusted” CFR early in the COVID-19 outbreak using an equivalent method with an assumed value of the time delay between onset and death (because the true value was not known): Yuan and colleagues [18] chose sample values of one, three and five days to give estimates from 3-13% for Italy in early March, while Wilson and colleagues [19] used 13 days to give 0.8-3.5% for China in early March.

### Application of TSD analysis to SARS 2003 outbreak in Hong Kong

To test the TSD analysis method in determining CFR in the middle of an outbreak, and compare to alternative methods, we analyse data from the SARS 2003 outbreak in Hong Kong (17 March to 11 July), obtained from the World Health Organization [20] and 3-day averaged. Figure 4 shows the cumulative and daily number of SARS cases and deaths in Hong Kong as a function of time.

**Figure 4:**
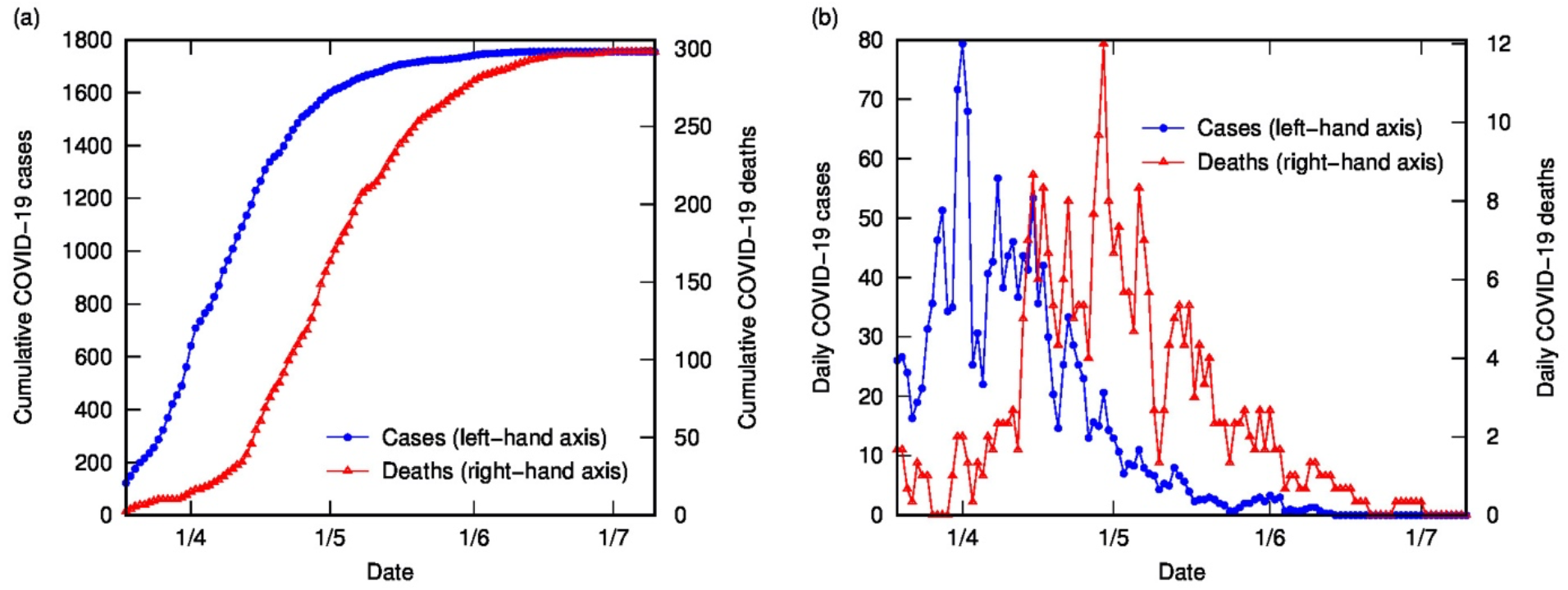
SARS cases and deaths in Hong Kong (2003), using 3-day averaged data: (a) Cumulative cases (left-hand axis) and deaths (right-hand axis); (b) Daily cases (left-hand axis) and deaths (right-hand axis).

It is apparent in Figure 4 that, as for COVID-19 data in Italy, the shapes of the distributions of cases and deaths are analogous. Time-shifted distribution analysis gives the following at the end of the outbreak: delay time is 22 days, and calculated CFR is 16.7%, close to the final crude CFR of 17.0%. The linear fit is reasonable given the noise in the data, as shown in Figure 5. If we perform TSD analysis serially over the course of the outbreak, reasonable estimates can be obtained from 17 April, giving values of 12-17% (with delay times of 17-22 days) converging on 16.7%, as shown in Figure 6. To compare, on 17 April the crude CFR is 5.3%, which is a significant underestimate of the true value. The delay time of 22 days is consistent with observations that the delay between onset and death for SARS is approximately three weeks [21]. We also applied TSD analysis to SARS data for other countries, giving a calculated CFR of 15% for Singapore and Canada (although data are noisy), and 13% for Taiwan.

**Figure 5:**
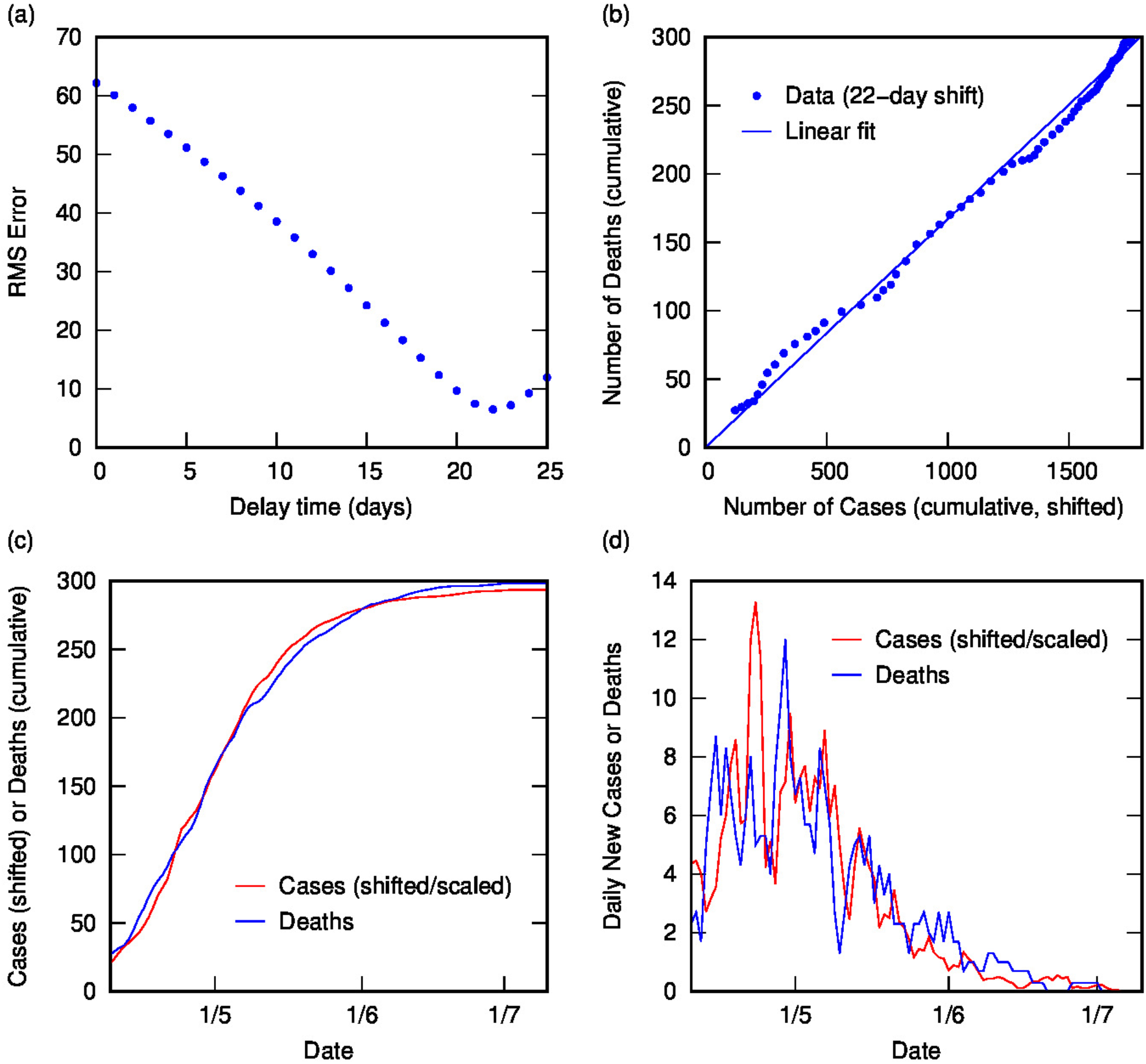
Time-shifted distribution analysis of SARS (2003) data for Hong Kong: (a) Root-mean-squared error in linear regression as a function of delay time, *t*_*d*_; (b) Linear regression for cumulative number of deaths as a function of cumulative number of cases (time-shifted by optimal *t*_*d*_); (c) Overlay of cumulative deaths and time-shifted (and scaled) cases as a function of time, using optimal *t*_*d*_; (d) Overlay of daily deaths and time-shifted (and scaled) cases as a function of time, using optimal *t*_*d*_.

**Figure 6:**
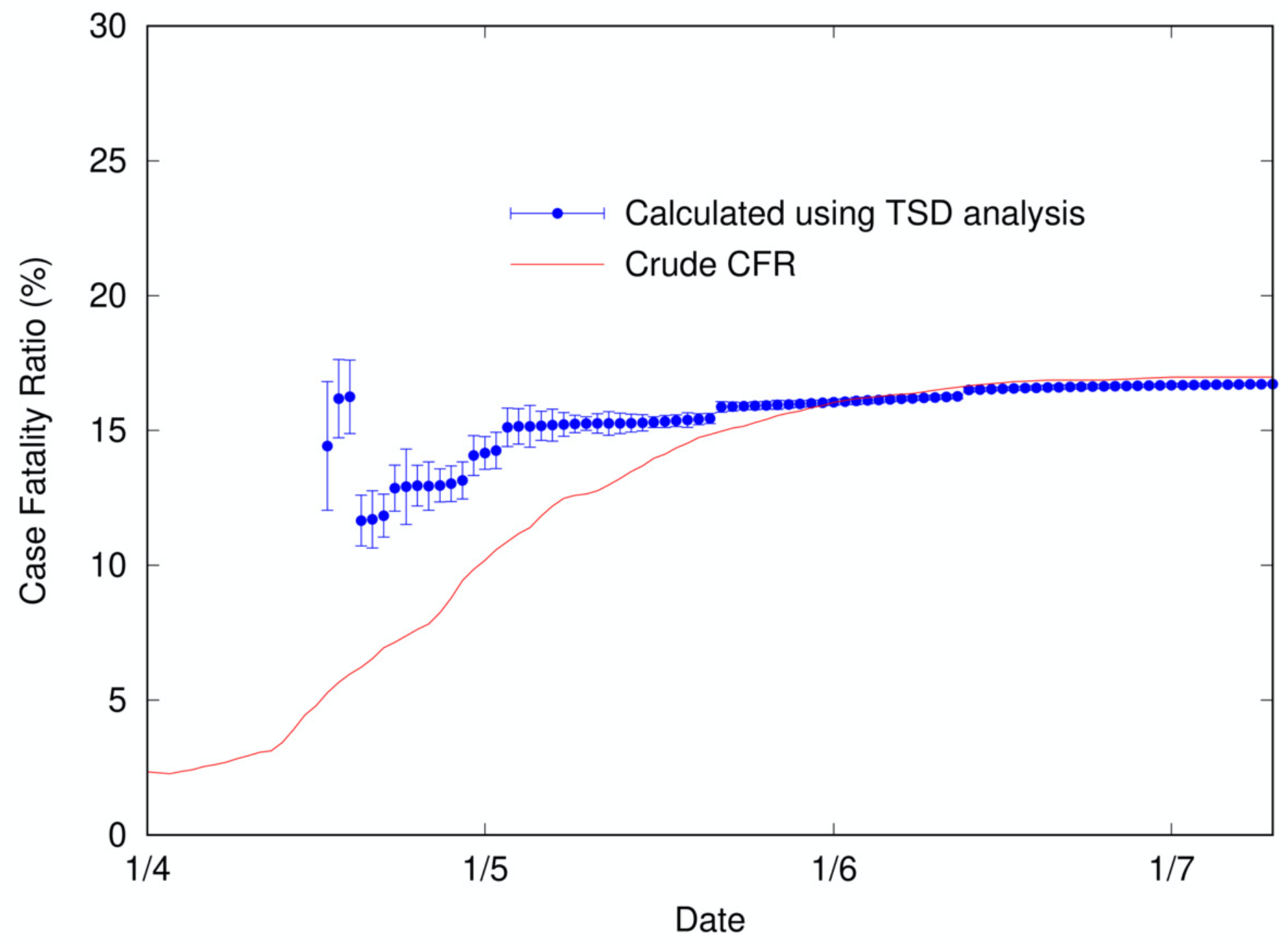
Calculated case fatality ratio (using TSD analysis) for SARS in Hong Kong (2003) as a function of time during an outbreak, alongside the crude CFR.

We can compare these estimates of the CFR with the more complex mathematical models of Nishiura and coworkers [22] and Ghani and coworkers [15] for the same SARS outbreak. The simple TSD analysis gives better predictions than both the parametric mixture model and modified Kaplan-Meier method described by Ghani et al. [15], which use individual case data(dates of hospitalisation and death or discharge from hospital) to estimate CFR using statistical methods. Such methods can provide earlier estimates (from 1 April, giving around 7-8% CFR) but are less accurate at this early stage than a simple estimate of CFR from data on closed cases (recoveries and deaths) at the same dates [15], and are later outperformed by our simple TSD method once sufficient data to perform the analysis are available. Further, TSD analysis requires only publicly reported case and death data (over time), which is easier to obtain than individual case data including onset dates.

Similarly, the model of Nishiura et al. [22] can provide much earlier estimates of CFR than our analysis but the accuracy of these estimates is uncertain and depends on the assumptions made. Their analysis requires data on the dates of onset of confirmed cases and the distribution of times from onset to death; the latter, in particular, is poorly known at the start of an outbreak of a new disease. Nishiura et al. [22] analyse the Hong Kong SARS data by assuming a simple exponential distribution for the time between onset and death, with a mean of 36 days (from Donnelly [21] for SARS cases in Hong Kong up to 28 April, although Donnelly used a gamma distribution), and using statistical sampling to predict the CFR. The fact that this model provides a reasonable prediction of CFR at a specific time (around the end of March) is likely fortuitous, given that it involves scaling the crude CFR by a constant factor and will therefore overestimate the CFR at later times (as well as very early times). Further, this method requires the use of parametrised data (the time distribution for onset to death) that are not available at the time that the predictions are purported to be made. In fact, when Nishiura et al. [22] apply the method to early H1N1 (swine flu) data in 2009 they are forced to use a time distribution calculated from historical data for H1N1 (Spanish) influenza from 1918-1919, which is problematic; a sensitivity analysis shows that the predicted CFR is sensitive to the choice of distribution parameters, making this method somewhat difficult to apply in the circumstances for which it is proposed.

In comparison, the time-shifted distribution analysis is both transparent and straightforward to implement, using only publicly available data and no assumptions, and can provide a reasonably early estimate (once exponential growth has sufficiently slowed) of CFR that converges to the “true” value. If the value of the time delay is approximately known early in the outbreak, this could be used to constrain the fitting procedure, but as observed already, it is difficult either to know the time delay between onset and death or to apply it to the time delay between reporting of cases and deaths.

### Time-shifted distribution analysis of international COVID-19 data

Time-shifted distribution analysis was performed on COVID-19 data from an extensive range of countries, using datasets from Johns Hopkins Center for Systems Science and Engineering [14], cross-checked and supplemented with data from Worldometers.com and 3-day averaged. For most countries (as for Italy), the analysis results in a robust linear fit and provides a stable estimate for CFR and delay time. These data are shown in Table 1, organised by region(Europe, Middle East, Asia, Oceania, North/Central America, South America, Africa) and then by CFR (decreasing). The corresponding plots of cases and deaths for each country are included in Appendix 1, to demonstrate the astonishing correlation between case and death time profiles over a huge range of locations and outbreak characteristics. For some few countries, either the data are insufficiently resolved (for example, still in exponential growth or very low numbers) or too noisy or unreliable for rigorous analysis. For other countries, the linear correlation is not robust and varies over time; notable examples are Sweden, Brazil and the USA, which are discussed in the following section. For countries listed in Table 1, most analyses use data up until the end of May, which is generally representative of the initial outbreak; for some countries with later outbreaks, later end dates are used. In many countries, more recent outbreaks have had dramatically different CFR values to initial outbreaks (due largely to improved testing rates); these can be analysed independently by selecting the time frame studied, but values presented here are for the initial outbreak in each country.

**Table 1:**
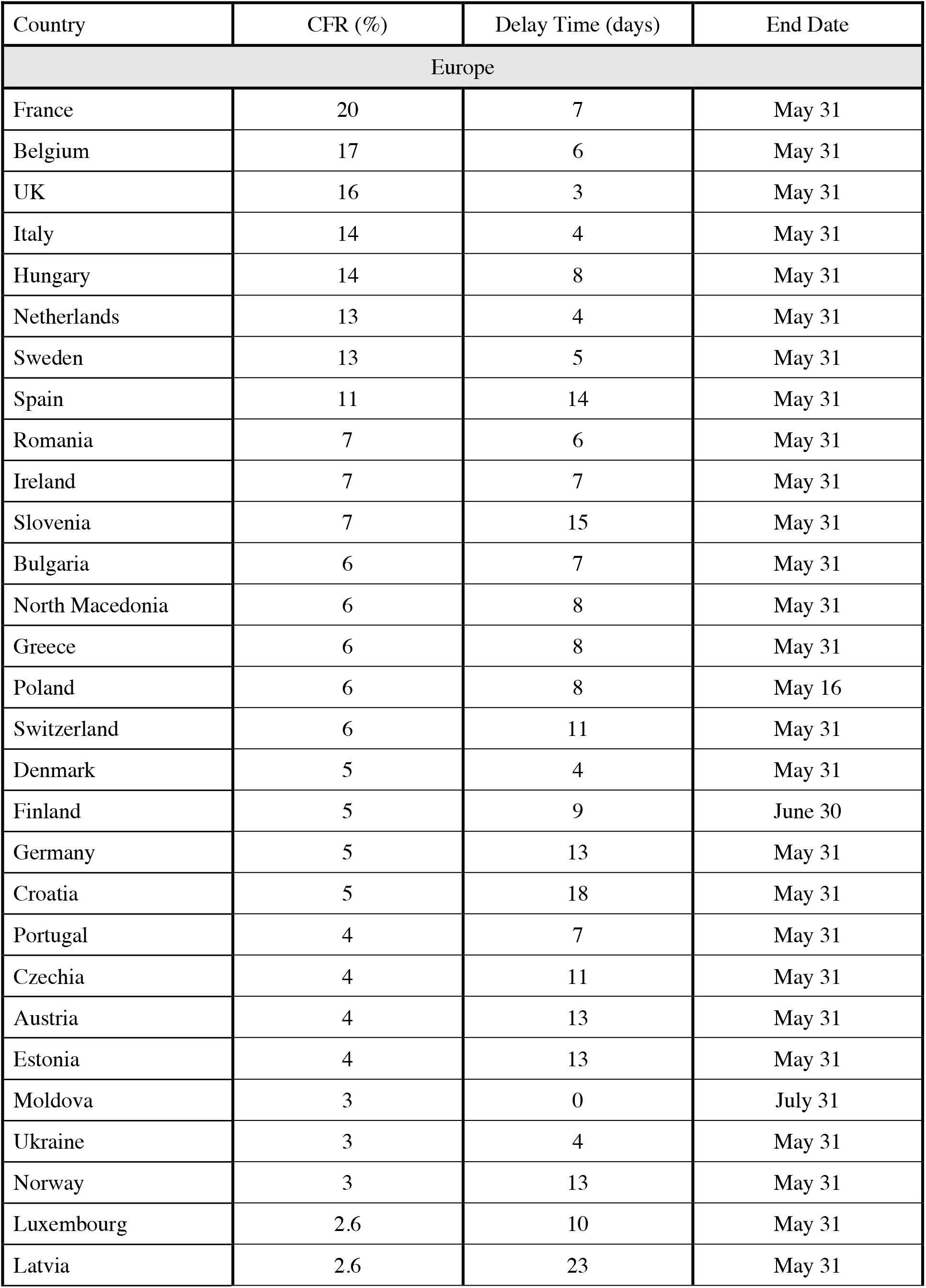
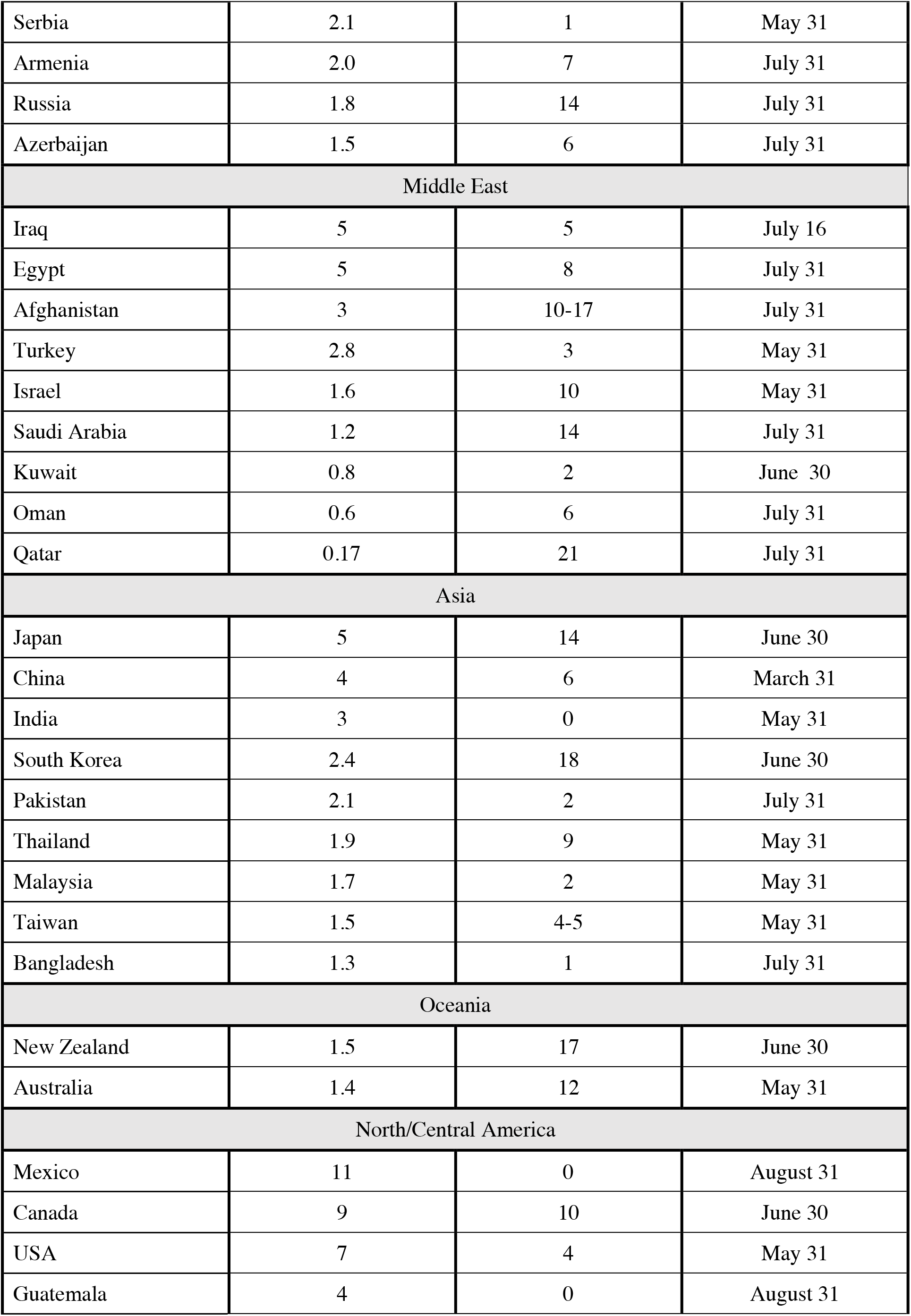
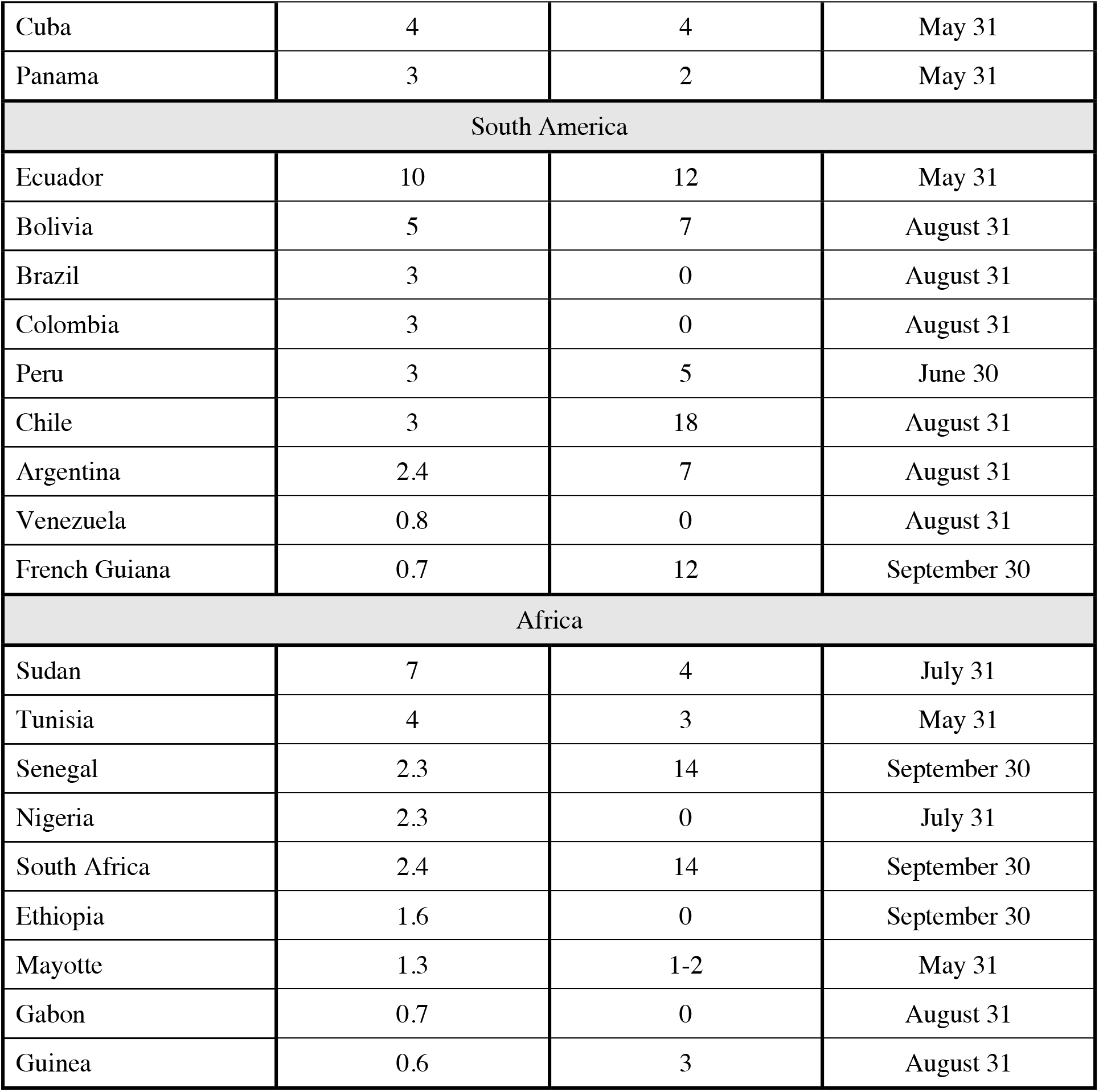
Case fatality ratio values and delay times calculated using time-shifted distribution analysis for a range of countries (initial outbreak), ordered by region and by CFR.

| Country | CFR (%) | Delay Time (days) | End Date |
| --- | --- | --- | --- |
| Europe |  |  |  |
| France | 20 | 7 | May 31 |
| Belgium | 17 | 6 | May 31 |
| UK | 16 | 3 | May 31 |
| Italy | 14 | 4 | May 31 |
| Hungary | 14 | 8 | May 31 |
| Netherlands | 13 | 4 | May 31 |
| Sweden | 13 | 5 | May 31 |
| Spain | 11 | 14 | May 31 |
| Romania | 7 | 6 | May 31 |
| Ireland | 7 | 7 | May 31 |
| Slovenia | 7 | 15 | May 31 |
| Bulgaria | 6 | 7 | May 31 |
| North Macedonia | 6 | 8 | May 31 |
| Greece | 6 | 8 | May 31 |
| Poland | 6 | 8 | May 16 |
| Switzerland | 6 | 11 | May 31 |
| Denmark | 5 | 4 | May 31 |
| Finland | 5 | 9 | June 30 |
| Germany | 5 | 13 | May 31 |
| Croatia | 5 | 18 | May 31 |
| Portugal | 4 | 7 | May 31 |
| Czechia | 4 | 11 | May 31 |
| Austria | 4 | 13 | May 31 |
| Estonia | 4 | 13 | May 31 |
| Moldova | 3 | 0 | July 31 |
| Ukraine | 3 | 4 | May 31 |
| Norway | 3 | 13 | May 31 |
| Luxembourg | 2.6 | 10 | May 31 |
| Latvia | 2.6 | 23 | May 31 |

Table 1 continued...
|  |  |  |  |
| --- | --- | --- | --- |
| Serbia | 2.1 | 1 | May 31 |
| Armenia | 2.0 | 7 | July 31 |
| Russia | 1.8 | 14 | July 31 |
| Azerbaijan | 1.5 | 6 | July 31 |
| Middle East |  |  |  |
| Iraq | 5 | 5 | July 16 |
| Egypt | 5 | 8 | July 31 |
| Afghanistan | 3 | 10-17 | July 31 |
| Turkey | 2.8 | 3 | May 31 |
| Israel | 1.6 | 10 | May 31 |
| Saudi Arabia | 1.2 | 14 | July 31 |
| Kuwait | 0.8 | 2 | June 30 |
| Oman | 0.6 | 6 | July 31 |
| Qatar | 0.17 | 21 | July 31 |
| Asia |  |  |  |
| Japan | 5 | 14 | June 30 |
| China | 4 | 6 | March 31 |
| India | 3 | 0 | May 31 |
| South Korea | 2.4 | 18 | June 30 |
| Pakistan | 2.1 | 2 | July 31 |
| Thailand | 1.9 | 9 | May 31 |
| Malaysia | 1.7 | 2 | May 31 |
| Taiwan | 1.5 | 4-5 | May 31 |
| Bangladesh | 1.3 | 1 | July 31 |
| Oceania |  |  |  |
| New Zealand | 1.5 | 17 | June 30 |
| Australia | 1.4 | 12 | May 31 |
| North/Central America |  |  |  |
| Mexico | 11 | 0 | August 31 |
| Canada | 9 | 10 | June 30 |
| USA | 7 | 4 | May 31 |
| Guatemala | 4 | 0 | August 31 |

Table 1 continued...
|  |  |  |  |
| --- | --- | --- | --- |
| Cuba | 4 | 4 | May 31 |
| Panama | 3 | 2 | May 31 |
| South America |  |  |  |
| Ecuador | 10 | 12 | May 31 |
| Bolivia | 5 | 7 | August 31 |
| Brazil | 3 | 0 | August 31 |
| Colombia | 3 | 0 | August 31 |
| Peru | 3 | 5 | June 30 |
| Chile | 3 | 18 | August 31 |
| Argentina | 2.4 | 7 | August 31 |
| Venezuela | 0.8 | 0 | August 31 |
| French Guiana | 0.7 | 12 | September 30 |
| Africa |  |  |  |
| Sudan | 7 | 4 | July 31 |
| Tunisia | 4 | 3 | May 31 |
| Senegal | 2.3 | 14 | September 30 |
| Nigeria | 2.3 | 0 | July 31 |
| South Africa | 2.4 | 14 | September 30 |
| Ethiopia | 1.6 | 0 | September 30 |
| Mayotte | 1.3 | 1-2 | May 31 |
| Gabon | 0.7 | 0 | August 31 |
| Guinea | 0.6 | 3 | August 31 |

The most notable result is the huge range in both delay times and calculated CFR estimates over different countries: from zero to 24 days’ delay and from less than one to 20% case fatality ratio. The highest ratios are calculated in Western Europe (up to 20%), followed by North America (up to 15%), South America (up to 10%), Africa (up to 7%), and lowest in the Middle East, Asia and Oceania (up to 5%). It is problematic to draw conclusions about relative COVID-19 virulence by comparing CFR values between countries, because of vast differences in testing and reporting regimes — in particular, the under-reporting of cases (including mild or asymptomatic cases) due to inadequate testing, but also differences in classification or recognition of COVID-19-related deaths. However, it is instructive to calculate in this way, for any given country, the proportion of detected cases that are currently proving fatal, for the purposes of public health management and planning. For comparison, Mazumder and colleagues [8] calculated case fatality ratios for a range of countries using recovery and death data from closed cases. They analysed eleven countries with high outcome rates and sufficient progression in the outbreak for analysis (at the end of April), but many of their calculated CFR values are much higher than our estimates — for example, estimated CFR above 30% for Italy, France and USA at the end of April — probably due to delays in recovery reporting, whereas estimated CFR values for Germany, China and South Korea match ours. The TSD analysis provides more reliable estimates for a broader range of countries, due to the greater availability of death and case data over recovery data.

The differences in delay times are also startling, ranging from zero to 24 days with no clear pattern. This delay between reported cases and deaths may be informative regarding the state of reporting or testing in a country but it is difficult to interpret. The mean delay between onset of symptoms and death has been estimated at 12-22 days using case data [2, 5, 7, 10, 12, 16], but there are also delays between onset of symptoms and testing, between testing and reporting of results, and in reporting of deaths. For example, in Sweden a mean delay of five days between onset of symptoms and the “statistical date” of a reported case (including one day from test to statistic) was reported [7]. In some countries, tests are only administered to the sickest patients (many days after onset), and in others, test results can take up to a few weeks. We note that for Australia and New Zealand, where case numbers have been low and testing extensive and rapid, the calculated time delay is more than ten days, whereas many of the harder-hit countries in western Europe and North America have much shorter calculated time delays.

Spain is an interesting case. Until 12 August, TSD analysis using Spanish data from the Worldometer website [23] gave a stable CFR of 10% with a delay time of one day. On that day, data were “adjusted retrospectively by national authorities: case counts adjusted from 2 February to 11 August and death counts adjusted from 26 April to 11 August” according to the World Health Organisation (WHO) [24]. Using the revised data, the TSD analysis provided an even more robust fit; the CFR was almost unchanged at 11% but the delay time was increased to 14 days. This means that early data from Spain, which were erratic, reflected a much shorter delay between reported cases and deaths. In fact, the death data were largely unaffected by the August revision, but the dates of reported cases had shifted nearly two weeks earlier, presumably to better capture the onset time. This shows that a short delay time can reflect late reporting of cases, due either to testing late in the progress of the disease (well after onset) or delays in providing test results (or both). This may explain the short delay times for the United Kingdom, Italy, the Netherlands and the USA, as well as many other countries (for example, zero days’ delay in Mexico). For countries that demonstrated reliable contact-tracing and testing regimes, such as Australia, New Zealand and Germany, delay times are close to 14 days, similar to the revised Spain data and consistent with the estimated time course of fatal disease.

An important conclusion from this analysis concerns the perils in calculating the CFR using established time distributions for onset to death obtained from case studies, as is common. The beauty of this simple method is its transparency — nothing is assumed and the data are enabled to speak for themselves, and can therefore give us information that we might not expect, rather than merely reflecting our assumptions.

## Discussion

We are interested not only in the versatility and simplicity of this method, but also in what conclusions may be drawn from the parameters calculated — namely, the CFR and the delay time. Reported CFR values for COVID-19 vary widely, but the best current estimates of the true infection fatality ratio or IFR (taking into account all infections including undiagnosed and asymptomatic) are around 0.6-0.7% [4, 6] based on cruise ship and population serology data. The very high CFR values calculated for many European countries in particular are probably vastly inflated due to the inadequate testing and overwhelmed health systems in these countries, which result in underestimation of case numbers. However, it is an oversimplification to assume that this is the only relevant factor that differs between countries, since we know that demographics and health systems (among other things) can also affect survival probability. Such an assumption has been used in various studies, in order to compare the effectiveness of different countries’ reporting systems and to correct case numbers [4, 25]. However, by assuming that the IFR is identical everywhere at all times, valuable information is lost and conclusions may be misleading.

In this study, we estimate the IFR from the CFR for a subset of countries using seroprevalence data (to correct case numbers) and excess death data (to correct death numbers). Along similar lines, Ioannidis [26] previously estimated the IFR for various countries using seroprevalence data and cumulative reported deaths at a corresponding date, although this does not account for either excess deaths or the relationship between cases and deaths over time; in fact, using seroprevalence and death data alone reintroduces the issue of the unknown time delay between cases and deaths, which must be approximated. We note that studies from very early in the pandemic provided initial estimates for the true prevalence of COVID-19 in specific places; a spatiotemporal transmission model applied to Wuhan [27] gave a prevalence factor of seven in January (published mid-March), and a statistical analysis study of testing data in the USA [28] estimated a prevalence factor of nine in April (published in May). These early prevalence studies can be useful in roughly correcting the CFR to estimate the IFR before rigorous seroprevalence data are available.

In Australia, case numbers have been generally low (especially before June) and testing rates high. It is unlikely that there have been appreciable unreported COVID-related deaths [29]. However, even with robust testing many cases will be undiagnosed, especially asymptomatic cases, which could constitute half of all infections [30]. A recent seroprevalence study of elective surgery patients in four states [31] estimated that the number of true infections was around 5-10 times the number of reported cases, although the authors state that the study cohort may not reflect the general population (older individuals overrepresented). This prevalence ratio gives an approximate IFR for Australia of 0.1-0.3%. Note that before June, most of Australia’s COVID-19 cases were returned travellers, which may affect the age distribution and baseline health of cases compared to the general population. New Zealand, Taiwan and Thailand are similarly circumstanced and have very similar CFR values, which are expected to reflect similar IFR values to Australia. Singapore, with its extremely low fatalities and extensive testing, did not return a robust result from TSD analysis; nonetheless, the crude CFR of 0.07% at the end of May is likely a lower bound for the IFR.

The USA is an interesting case study. The TSD analysis is problematic because the relationship between cases and deaths changes over time, causing mismatch between case and death distributions and a downward drift in both CFR and delay time. This may be due to incomplete data, or changes in testing or reporting over time, which can affect both delay time and case numbers. Alternatively, the case fatality ratio may be truly changing over time, due to changes in treatment approach or in the demographics (or location) of COVID-19 cases [32]. In the USA, there is also heterogeneity between states. To demonstrate, we present the time-shifted distribution analysis for the USA in Figure 7, and for the state of New Jersey (which has the highest mortality rate in the USA) in Figure 8. For the USA as a whole, there is clear variation over time in the relationship between time-shifted case and death data, demonstrated in both the poor linear fit and the mismatch in distribution profiles. If we scrutinise individual state data, some US states (including New Jersey, Illinois, Massachusetts, New Mexico, Ohio and Pennsylvania) manifest a very reliable TSD analysis, but others do not (for example, California, North Carolina, Oklahoma and Texas). Data from New Jersey (Figure 8) give a stable CFR around 8-9%, comparable to New York, Massachusetts and Pennsylvania, while Ohio gives 7% and Illinois and New Mexico give 5%.

**Figure 7:**
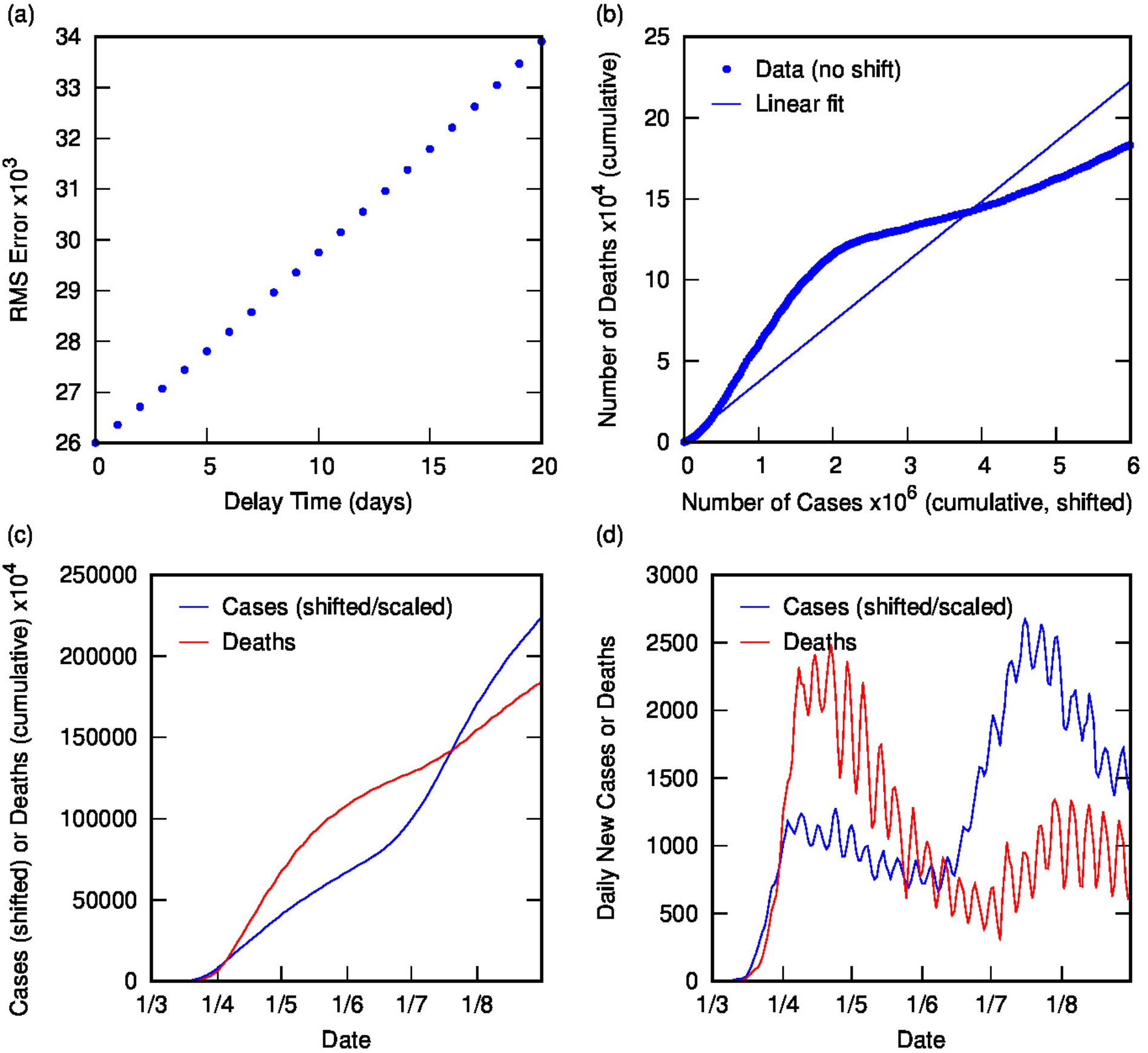
Time-shifted distribution analysis for USA at the end of August: (a) Root-mean-squared error in linear regression as a function of delay time, *t*_*d*_; (b) Linear regression for cumulative number of deaths as a function of cumulative number of cases (time-shifted by optimal *t*_*d*_); (c) Overlay of cumulative deaths and time-shifted (and scaled) cases as a function of time, using optimal *t*_*d*_; (d) Overlay of daily deaths and time-shifted (and scaled) cases as a function of time, using optimal *t*_*d*_. Note the mismatch between distributions of death and cases.

**Figure 8:**
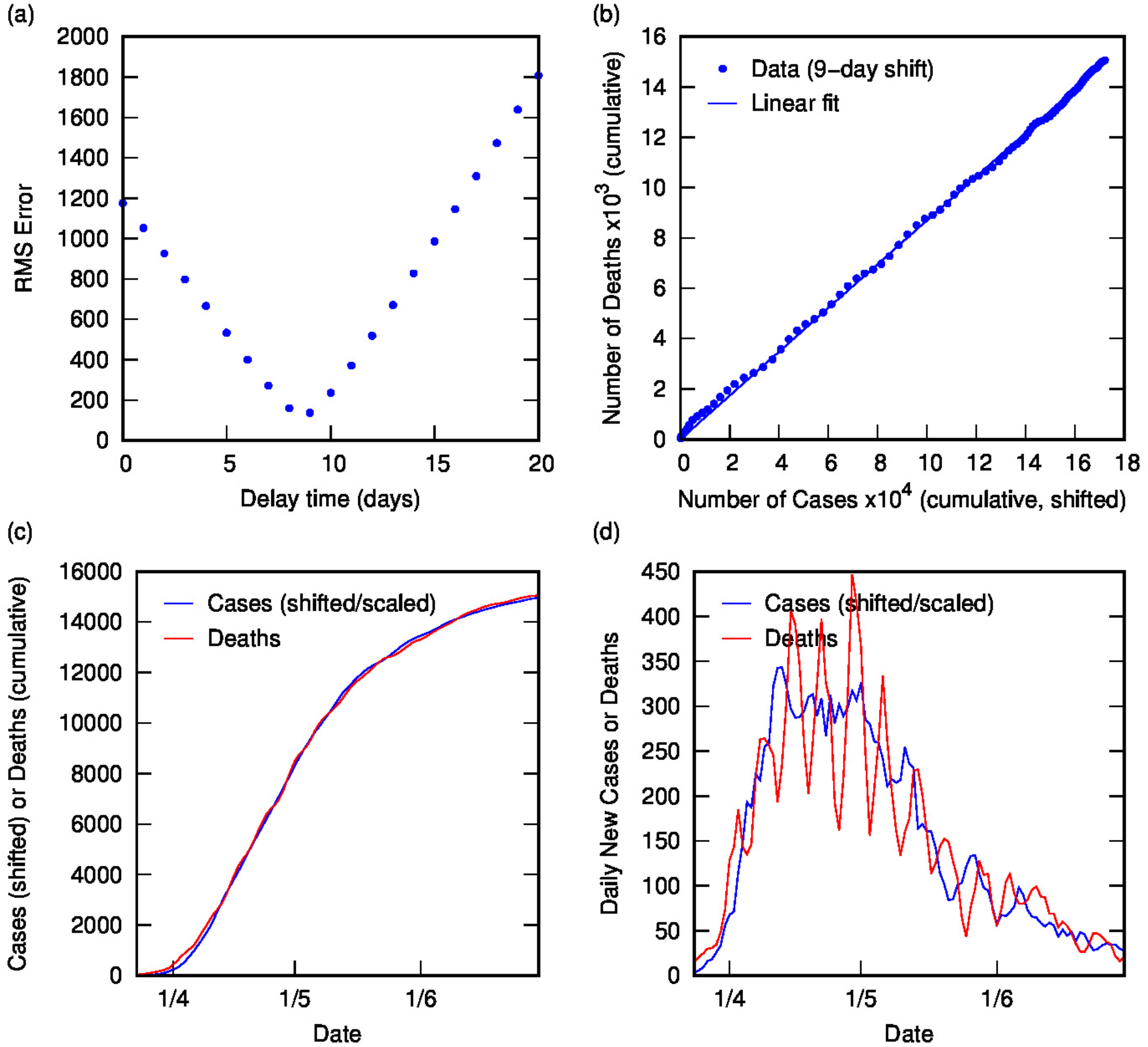
Time-shifted distribution analysis for New Jersey: (a) Root-mean-squared error in linear regression as a function of delay time, *t*_*d*_; (b) Linear regression for cumulative number of deaths as a function of cumulative number of cases (time-shifted by optimal *t*_*d*_); (c) Overlay of cumulative deaths and time-shifted (and scaled) cases as a function of time, using optimal *t*_*d*_;(d) Overlay of daily deaths and time-shifted (and scaled) cases as a function of time, using optimal *t*_*d*_.

One potential reason for the mismatch of case and death data in the USA as a whole (and many of its states) is the under-reporting of cases due to the low level of testing, which varies over time. One measure of the adequacy of testing is the share of daily COVID-19 tests that return a positive result, known as the positive test rate (PTR). The WHO has suggested a PTR around 3-12% (or less) as a benchmark of adequate testing [33]. In the USA, the positive test rate reached maximum levels in April, with values between 18 and 22% from 1-21 April [33], which is the region of greatest discrepancy between case and death profiles in the initial outbreak, as seen in Figure 7. We would expect that such a high PTR indicates that case numbers during this time are greatly underestimated, which may explain the poor fit from TSD analysis and the high CFR. Similar effects are seen in data from Sweden and Brazil, which also had low and variable testing rates and high PTR. Recent seroprevalence studies in many states of the USA from March to May [34] suggest that there were at least eleven times as many infections as reported cases before the end of May. Excess death data indicate that COVID-related deaths may be higher than reported by a factor of 1.4 for the same period [35]. Using these correction factors for the CFR, the estimated IFR for the USA is 1.0% or below. For comparison, a Worldometers calculation estimated an IFR of 1.4% in New York City in May [23], using a prevalence ratio of ten from an early antibody study [36, 37].

In Europe, many of the most affected countries have very high case fatality ratios, often combined with relatively short delay times. For some of these countries, seroprevalence studies provide estimates of the degree of undercounting of cases during the initial outbreak [38-46], which can be utilised along with excess death data [35] to estimate the infection fatality ratio. These IFR values are shown in Table 2 along with the correction factors used. Some of the seroprevalence data is preliminary, including studies of Germany, Sweden and Italy, and others are for specific regions of the country and may not be representative. Nonetheless, the estimated IFR values are reasonable: Switzerland and Germany are around 0.6%, above Australia and below Sweden and USA at around 0.8%; Belgium, UK and Spain are between one and two percent; and Italy higher at around 3%. Ioannidis [26] also calculated the IFR for many of these countries using seroprevalence studies, but using only single-time seroprevalence and death data with an assumed delay time (generally a week after the midpoint of the seroprevalence survey); these are also shown in Table 2 and are broadly consistent with our values except where excess deaths are significant (e.g. Spain). Our value for Germany is somewhat higher but we expect that it is more reliable, using the scaling factor for cases [38] with our calculated CFR rather than the absolute number of deaths at a certain date in the German town of Gangelt [26], which is very low and reflects a date early in the German outbreak.

**Table 2:** Estimated IFR from CFR (calculated in this work), using scaling factors from seroprevalence and excess death data.

| Country | CFR<br>[this work] | Scaling factors |  | IFR estimate<br>[this work] | IFR from<br>Ioannidis<sup>1</sup> [26] |
| --- | --- | --- | --- | --- | --- |
|  |  | Deaths<br>(excess) | Cases<br>(prevalence) |  |  |
| Australia | 1.4 | 1.0 | 5-10 [31] | 0.1-0.3 |  |
| Switzerland | 6 | 1.0 | 10-12 [39] | 0.5-0.6 | 0.45 |
| Germany | 5 | 0.8 | 5-7 [38] | 0.5-0.8 | 0.28 |
| Sweden | 13 | 1.2 | 17-21 [40] | 0.7-0.9 | 0.71 |
| USA<sup>2</sup> | 7 | 1.4 | 9-13 [34] | 0.7-1.0 | 0.65 |
| Belgium | 17 | 1.0 | 13-15 [42] | 1.1-1.3 | 1.09 |
| UK | 15 | 1.3 | 14-15 [41] | 1.3-1.4 |  |
| Spain | 11 | 1.5 | 9-12 [43, 44] | 1.3-1.8 | 1.15 |
| Italy | 14 | 1.5 | 6-7 [45, 46] | 3.0-3.5 |  |

<sup>1</sup> IFR from Ioannidis [26] uses seroprevalence and concomitant deaths at a single time point.
<sup>2</sup> USA value is a weighted mean of six states.

Although the calculated IFR values are only approximate and subject to revision, it is conceivable that higher IFR values may reflect higher fatality ratios in particular places at particular times, due to overwhelmed health systems in hard-hit areas or specific demographics or baseline health of affected populations. For example, it is reasonable to conclude that in Lombardy, Italy, the older population and overwhelmed health system caused a higher fatality ratio compared to other places. In fact, the difference in age distribution of cases between Italy and Australia up to the end of May (using data from [47] and [48]) can alone account for a factor of three in the IFR. Therefore, while differences in testing and reporting between different countries undoubtedly account for much of the variation in IFR between countries, we neither expect nor find that IFR is the same for all COVID-19 outbreaks. Country-specific factors that influence IFR and differ between countries include testing and reporting, age demographics [49], health-care systems and treatments [32], mask-wearing and other behaviours, climate and culture, transport infrastructure and community mobility [50], genetic factors or prevalence of particular antibodies that affect immune response [51].

There is some evidence that the IFR might be decreasing over time in some countries, especially those experiencing a “second wave”. This is observed, for example, in the data for the USA in Figure 7, demonstrated in the increasing mismatch in case and death distributions later in the outbreak. We can use the TSD analysis to analyse the latter part of the outbreak (from July to September), giving a CFR of 1.5-1.7% and a delay time of 2-3 weeks. Similar analyses for individual states of the USA give stable CFR values from 1.1-2.3% with delay times between four and 24 days, with a mean of 1.6% CFR and 17 days’ delay over states with robust fits. This later CFR is far lower than the value of 7% calculated early in the outbreak. We observe similar effects in various other countries post-July including Japan (reduced to 1.1% and 22 days’ delay) and Spain, France and Portugal (all reduced to 0.8-1.3%, 12-29 days’ delay). These values are all similar and may reflect a reasonable estimate for CFR when testing is adequate; we would still expect the IFR to be lower by a factor of at least two due to undiagnosed and asymptomatic cases. A decrease in CFR over time may also indicate a change in the demographics of the case load or improvements in treatment or even an increasing time delay between reported cases and deaths, perhaps due to earlier diagnosis.

We also find that, in countries where the time delay is significant, the TSD analysis can serve in a predictive capacity for numbers of deaths, using the linear relationship between deaths and time-shifted cases. Figure 9 shows this prediction for the second phase of the COVID-19 outbreak in France from August. Using parameters calculated from TSD analysis for August to mid-October, reported case data can be time-shifted and linearly scaled to predict daily deaths for France for the next three weeks. This is useful for public health planning, as well as decision-making regarding implementation of restrictions.

**Figure 9:**
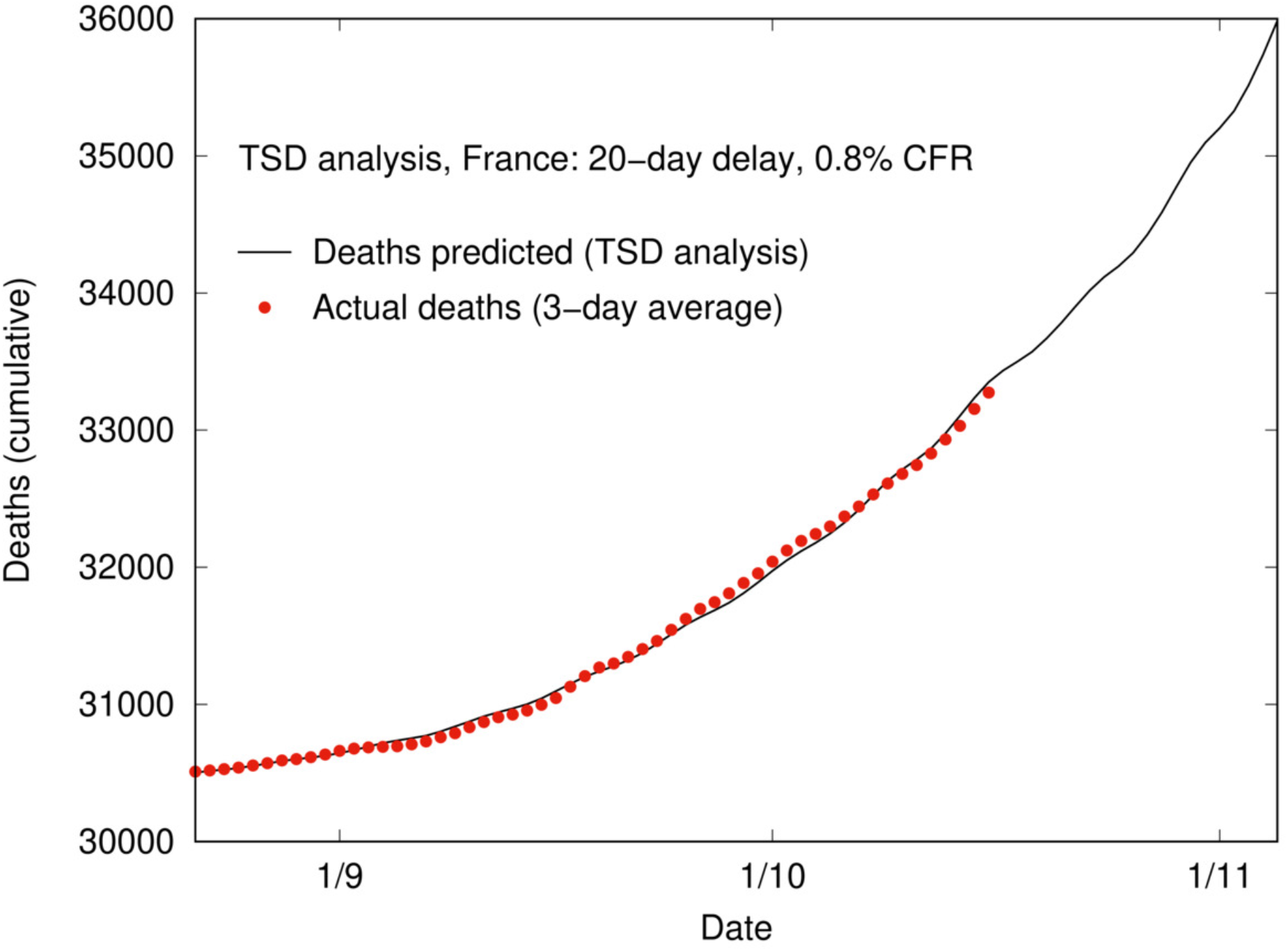
Using TSD analysis to predict deaths over time based on case data, delay time and CFR in France from August. Prediction uses 3-day averaged case data up to 16 October, shifted (20 days) and linearly scaled using the CFR (0.8%).

## Conclusion

The time-shifted distribution analysis is a straightforward way to predict CFR over time, using only publicly available data on cases and deaths and requiring no assumptions or parametrisations regarding the progress of the illness. The beauty of this method is in its transparency and simplicity; the lack of assumptions allows more to be gained from the data, including trends that may be unexpected or changing over time. This analysis method has particular utility early in an outbreak, once sufficient data are available for a robust fit (beyond the exponential growth phase). Without the benefit of hindsight, the TSD-calculated values for CFR and time delay between cases and deaths can shed light on the virulence of a disease and on the conditions that a particular country may be facing. Excess death data (where available) may be used to correct death data, while positive test rates and other indicators or models of testing adequacy can often give an early rough idea of the true prevalence relative to reported case numbers. These data can be used to interpret the CFR calculated using TSD analysis early in an outbreak, and to approximate the IFR.

Our estimates of IFR range from 0.3-3%, with higher values observed for countries that experienced more severe outbreaks, perhaps reflecting the negative influence of overwhelmed health systems and the spread of disease to more vulnerable populations. The calculated time delay is also potentially informative; for example, the one-day delay calculated from early data in Spain reflects the breakdown of testing and reporting systems at that time, whereas the revised delay time of 14 days shows the recovery of the system and the likely delay between case diagnosis and death. In this way, TSD analysis of data from a particular place at a particular time can give useful local information on the progression of an outbreak to inform public health planning and policy.

## Data Availability

The data used this study are publicly available at the links below.

https://github.com/pomber/covid19

https://www.worldometers.info/coronavirus/

https://github.com/imdevskp/sars-2003-outbreak-data-webscraping-code

https://github.com/TheEconomist/covid-19-excess-deaths-tracker

https://ourworldindata.org/coronavirus-testing

## Acknowledgements

We thank Dr Nick Golding (Curtin University) for many helpful conversations and comments on the manuscript.

## Data Availability Statement

The data used this study are publicly available. COVID-19 data are from the COVID-19 Data Repository by the Center for Systems Science and Engineering (CSSE) at Johns Hopkins University at https://github.com/CSSEGISandData/COVID-19 (via https://github.com/pomber/covid19), and from Worldometer at https://www.worldometers.info/coronavirus/. SARS data are from the World Health Organization at https://www.who.int/csr/sars/country/en/ (via https://www.kaggle.com/imdevskp/sars-outbreak-2003-complete-dataset). Excess death data are from the Economist’s COVID-19 excess deaths tracker repository at https://github.com/TheEconomist/covid-19-excess-deaths-tracker, and positive COVID-19 test rates are from Our World in Data at https://ourworldindata.org/coronavirus-testing.

