## Appendix 1 for "Estimating the Case Fatality Ratio for COVID-19 using a Time-Shifted Distribution Analysis"

### Overlay of daily COVID-19 cases (time-shifted and scaled) and deaths for many countries

Time-shifted distribution analysis gives two parameters: the time delay between case and death distributions and the case fatality risk, which acts as a linear scaling factor between time-shifted cases and deaths. The following plots overlay the daily time-shifted and scaled case distribution with the daily death distribution for each of the countries listed in Table 1, demonstrating excellent correlations over diverse outbreak characteristics, even where data are noisy.

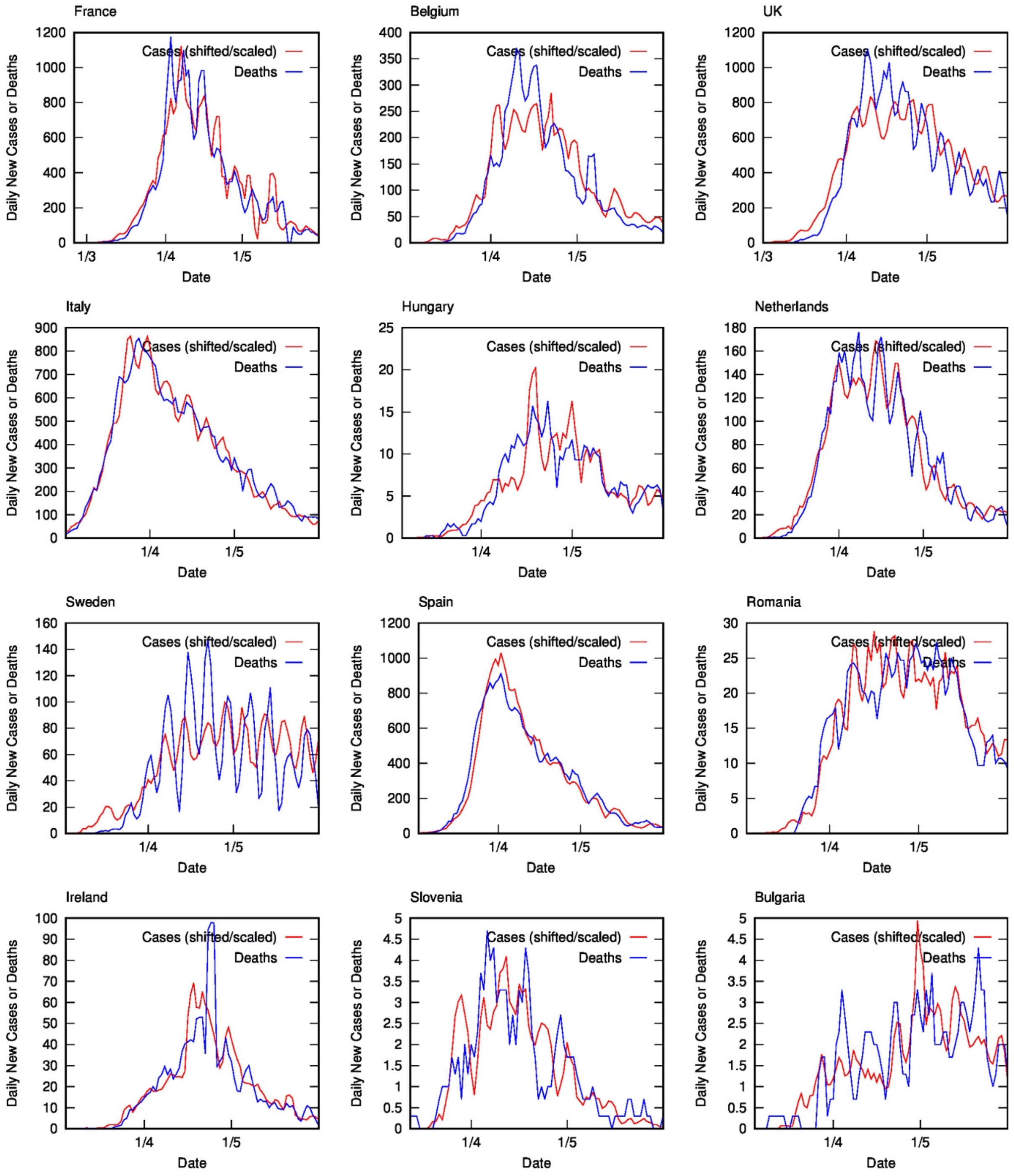

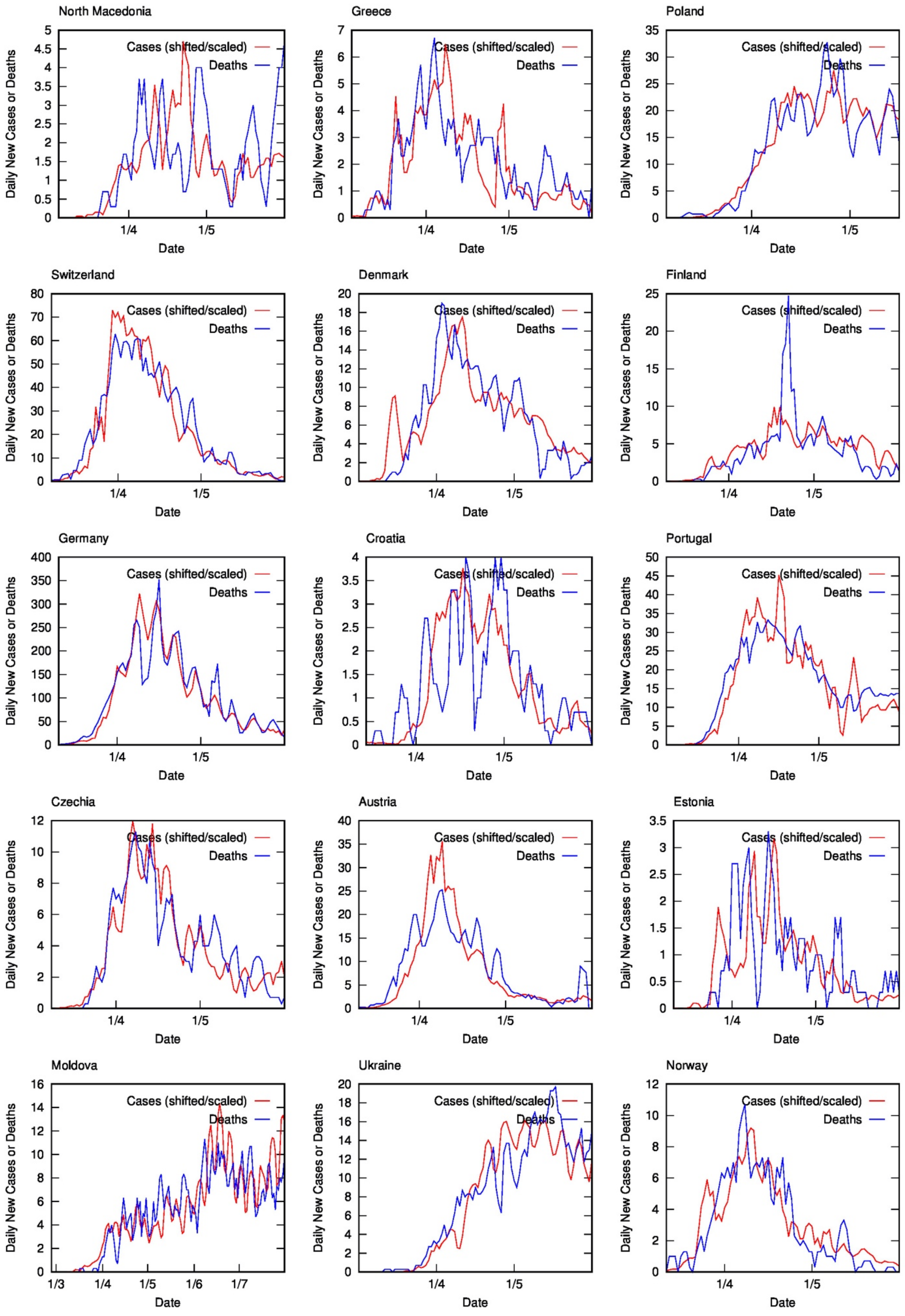
North Macedonia
Greece
Poland
Switzerland
Denmark
Finland
Germany
Croatia
Portugal
Czechia
Austria
Estonia
Moldova
Ukraine
Norway
Cases (shifted/scaled)
Deaths
Daily New Cases or Deaths
Date
1/3
1/4
1/5
1/6
1/7

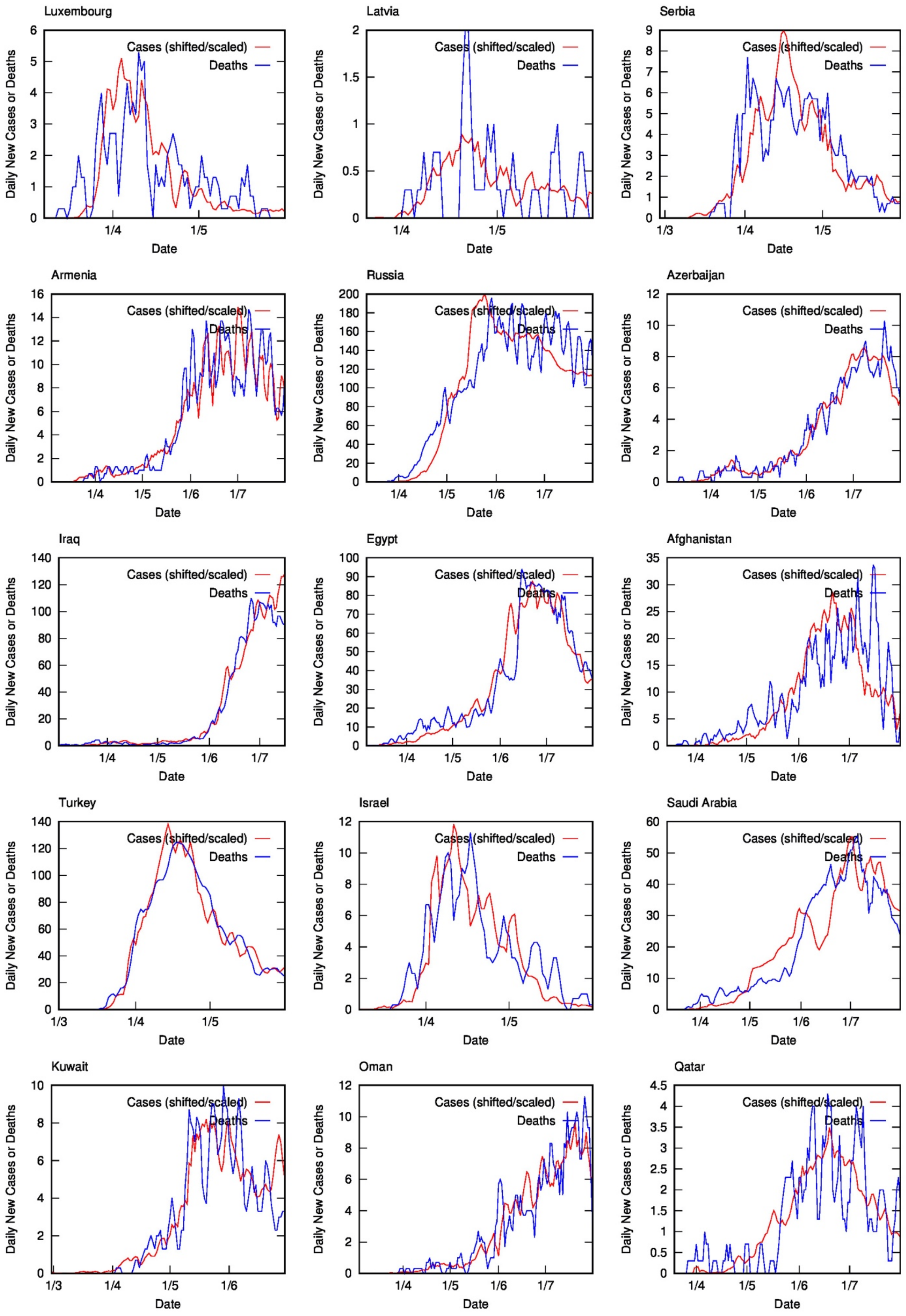
Luxembourg
Latvia
Serbia
Armenia
Russia
Azerbaijan
Iraq
Egypt
Afghanistan
Turkey
Israel
Saudi Arabia
Kuwait
Oman
Qatar
Cases (shifted/scaled)
Deaths
Daily New Cases or Deaths
Date

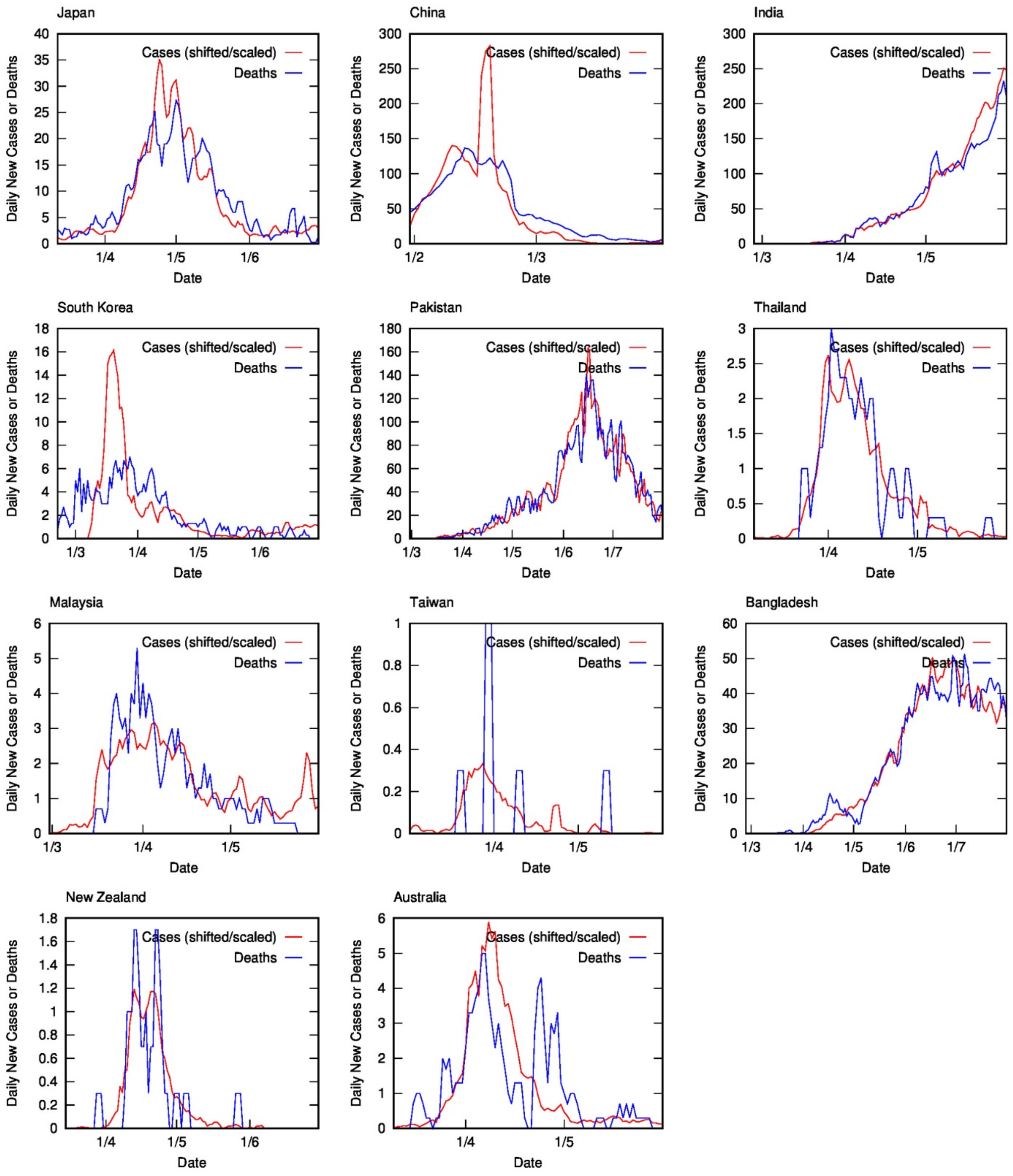
Japan
Cases (shifted/scaled)
Deaths
Daily New Cases or Deaths
Date
China
Cases (shifted/scaled)
Deaths
Daily New Cases or Deaths
Date
India
Cases (shifted/scaled)
Deaths
Daily New Cases or Deaths
Date
South Korea
Cases (shifted/scaled)
Deaths
Daily New Cases or Deaths
Date
Pakistan
Cases (shifted/scaled)
Deaths
Daily New Cases or Deaths
Date
Thailand
Cases (shifted/scaled)
Deaths
Daily New Cases or Deaths
Date
Malaysia
Cases (shifted/scaled)
Deaths
Daily New Cases or Deaths
Date
Taiwan
Cases (shifted/scaled)
Deaths
Daily New Cases or Deaths
Date
Bangladesh
Cases (shifted/scaled)
Deaths
Daily New Cases or Deaths
Date
New Zealand
Cases (shifted/scaled)
Deaths
Daily New Cases or Deaths
Date
Australia
Cases (shifted/scaled)
Deaths
Daily New Cases or Deaths
Date

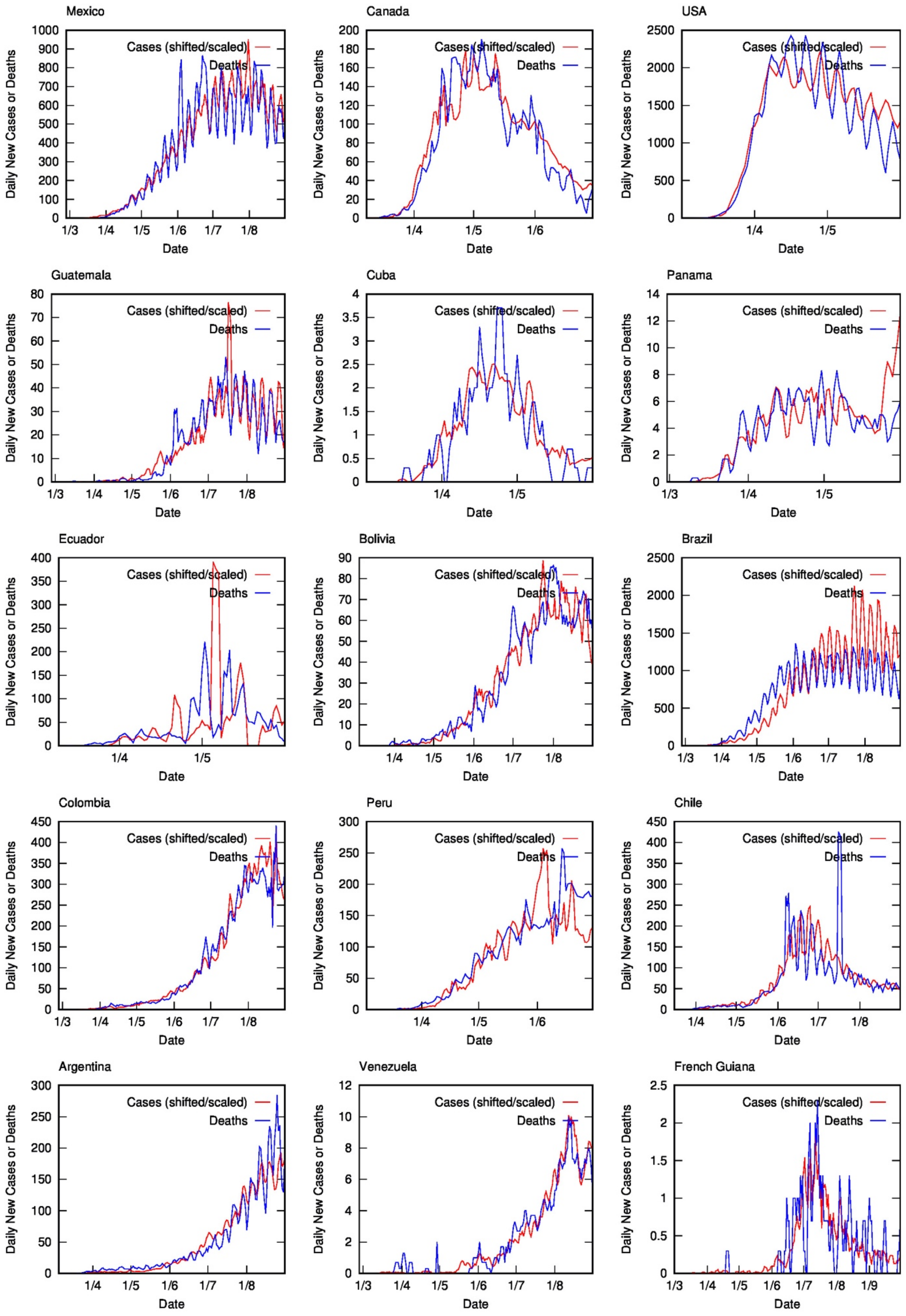
Mexico
Canada
USA
Guatemala
Cuba
Panama
Ecuador
Bolivia
Brazil
Colombia
Peru
Chile
Argentina
Venezuela
French Guiana
Cases (shifted/scaled)
Deaths
Daily New Cases or Deaths
Date

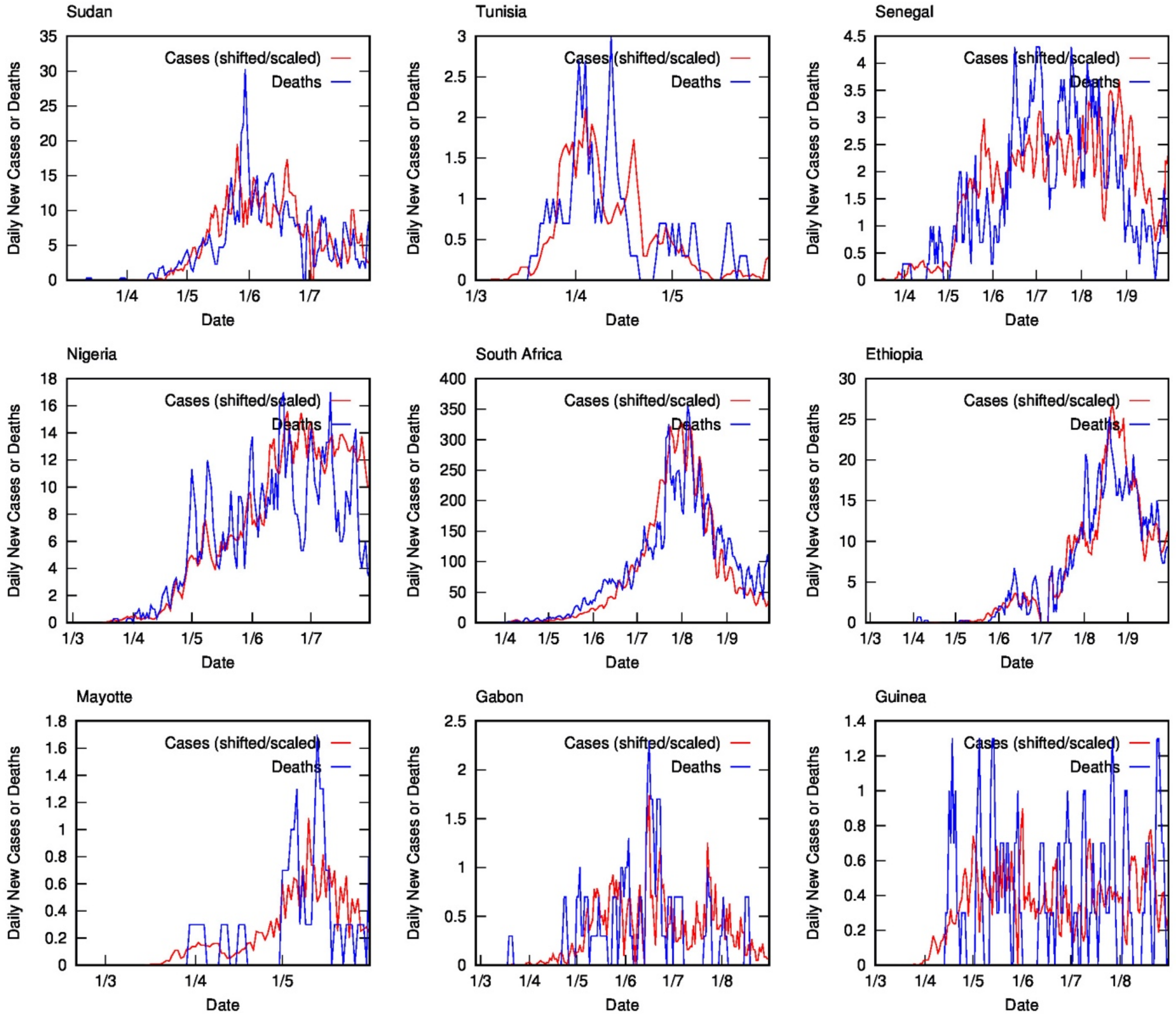
Sudan
Cases (shifted/scaled)
Deaths
Daily New Cases or Deaths
35
30
25
20
15
10
5
0
1/4
1/5
1/6
1/7
Date
Tunisia
Cases (shifted/scaled)
Deaths
Daily New Cases or Deaths
3
2.5
2
1.5
1
0.5
0
1/3
1/4
1/5
Date
Senegal
Cases (shifted/scaled)
Deaths
Daily New Cases or Deaths
4.5
4
3.5
3
2.5
2
1.5
1
0.5
0
1/4
1/5
1/6
1/7
1/8
1/9
Date
Nigeria
Cases (shifted/scaled)
Deaths
Daily New Cases or Deaths
18
16
14
12
10
8
6
4
2
0
1/3
1/4
1/5
1/6
1/7
Date
South Africa
Cases (shifted/scaled)
Deaths
Daily New Cases or Deaths
400
350
300
250
200
150
100
50
0
1/4
1/5
1/6
1/7
1/8
1/9
Date
Ethiopia
Cases (shifted/scaled)
Deaths
Daily New Cases or Deaths
30
25
20
15
10
5
0
1/3
1/4
1/5
1/6
1/7
1/8
1/9
Date
Mayotte
Cases (shifted/scaled)
Deaths
Daily New Cases or Deaths
1.8
1.6
1.4
1.2
1
0.8
0.6
0.4
0.2
0
1/3
1/4
1/5
Date
Gabon
Cases (shifted/scaled)
Deaths
Daily New Cases or Deaths
2.5
2
1.5
1
0.5
0
1/3
1/4
1/5
1/6
1/7
1/8
Date
Guinea
Cases (shifted/scaled)
Deaths
Daily New Cases or Deaths
1.4
1.2
1
0.8
0.6
0.4
0.2
0
1/3
1/4
1/5
1/6
1/7
1/8
Date
